# Identifiability and predictability of integer- and fractional-order epidemiological models using physics-informed neural networks

**DOI:** 10.1101/2021.04.05.21254919

**Authors:** Ehsan Kharazmi, Min Cai, Xiaoning Zheng, Guang Lin, George Em Karniadakis

## Abstract

We analyze a plurality of epidemiological models through the lens of physics-informed neural networks (PINNs) that enable us to identify multiple time-dependent parameters and to discover new data-driven fractional differential operators. In particular, we consider several variations of the classical susceptible-infectious-removed (SIR) model by introducing more compartments and delay in the dynamics described by integer-order, fractional-order, and time-delay models. We report the results for the spread of COVID-19 in New York City, Rhode Island and Michigan states, and Italy, by simultaneously inferring the unknown parameters and the unobserved dynamics. For integer-order and time-delay models, we fit the available data by identifying time-dependent parameters, which are represented by neural networks (NNs). In contrast, for fractional differential models, we fit the data by determining different time-dependent derivative orders for each compartment, which we represent by NNs. We investigate the identifiability of these unknown functions for different datasets, and quantify the uncertainty associated with NNs and with control measures in forecasting the pandemic.

---

Since the outbreak of the novel coronavirus at the beginning of 2020, over 100,000 papers have been published related to COVID-19, with a significant portion of them focusing on epidemiological models that describe the observed and unobserved dynamics, primarily in postdictive mode although some models are also used for short-term forecasting. These models represent lumped dynamics of a big city, a state or a country but suffer from large uncertainties^1^, resulting primarily from the lack of identifiability as well as the noise in the available sparse data. This lack of idetifiability is related to modeling assumptions (structural identifiability), data availability, and complex biological characteristics of virus transmission, which are hard to measure, they remain largely unknown^2^, and include various emerging mutations of the virus^3^. The true scientific challenge is to recognize the large limitations of these (potentially useful) models, identify the multiple sources of uncertainty, and suggest new flexible models that can deal with seasonal variation in susceptibility, time-delays, noisy data, under-determined systems, non-Markovian behavior, and inherent stochasticity^4^. In addition to seasonal variation in transmission, e.g., due to weather or mobility, for a given model the data uncertainty is usually propagated into model parameters, rendering them as random variables/processes with an underlying probability distribution. The uncertainties in the input parameters affect the model predictability adversely rendering many of these models inadequate for any decision making, as they lack robustness, which is a measure of the extent to which the forward solvers amplify uncertainties from the input to the output^5^. In general, quantification of parametric input uncertainty is only based on a single given model, hence ignoring the bigger source of uncertainty associated with the model structure. A clear example of uncertainty associated with several different models in analyzing and predicting the dynamics of this complex system can be found in the COVID-19 Forecast Hub^6^, which is a public online server that serves as a central repository of forecasts and predictions from over 50 international research groups. Even with a 95% confidence interval, one can see that the prediction of all of these different models can vary drastically (see a snapshot of vastly different predictions in Supplementary Section 2).

Parameter identifiability relates to model uncertainty for a specific model but here we want to pose questions beyond this uncertainty notion, and aim to investigate two fundamentally different approaches to epidimiological modeling. In the classical approach, the models are obtained by lumping the complex system into several compartments governed by a (nonlinear coupled) system of ordinary differential equations (ODEs), where we adjust the (possibly time-dependent) parameters of the model to fit the available data. This is of course all we can do because of the constraint of using pre-fabricated integer-order differential operators that represent fixed rate dynamics at all times. A variation of this is to introduce time-delays into the ODEs to account for some memory effects. An opposite way of constructing a model is to tweak the differential operators instead of tweaking the parameters, using, for example, fractional derivatives with their orders (and hence the rate of dynamics) determined directly by the data, while we use simple parameters with some nominal values in front of the operators. By considering these two classes, we expand both the identifiability and predictability analyses in a broader perspective that allows us to investigate a plurality of epidemiological models and test their performance with diverse COVID-19 datasets for cities, states, and countries. In particular, we consider several different integer-order, fractional-order and time-delay models, each expressed as a system of ODEs, where the parameters are either biologically determined and fixed or time-dependent. The integer-order and time-delay models fall into the first class of modeling with time-dependent parameters and unknown time-delay duration to fit the data. The fractional-order models are the new paradigm of modeling, where we tune the time-dependent operator to fit the available data.

In this broader framework, where the unknowns to be determined in the integer-order and time-delay models or the derivative orders in the fractional-order models are functions of time, the inverse problem to solve is particularly challenging. Several numerical methods have been developed to solve the inverse problem of inferring model (mostly constant) parameters from available data. They typically convert the problem into an optimization problem, and then, formulate a suitable estimator by minimizing an objective function, see e.g.^7–15^. Here, we overcome this difficulty by developing physics-informed neural networks (PINNs) both for integer-order and fractional-order models^16^. PINNs provide a flexible computational tool that encodes the ODEs into the neural networks to satisfy the equations while accurately fitting the data. In other words, parameter inference and simulation of the observed and unobserved dynamics take place simultaneously. The PINN formulation allows the parameters to be time-dependent functions, represented by separate neural networks, and this makes the inverse problem of inferring the parameters and unobserved dynamics from available data readily applicable to several different models. The structural and practical identifiability of integer-order models have been recently studied in^17^. However, this analysis cannot be applied to the fractional models or even to integer-order models with parameters, which are continuous functions of time. The unique determination of the derivatives’ variable order in the case of a time-fractional linear diffusion equation on a general multi-dimensional domain was proved in^18^. To the best of our knowledge, existence and uniqueness of the solution have not been proven for the inverse problem of identifying variable order for the nonlinear system of equations. Through systematic numerical experiments, in this paper we study and discuss the identifiability of different time-dependent fractional-orders of fractional models by using different amounts of data. If limited to only the reported available data, the inference is not very robust and the inferred fractional orders may exhibit relatively large uncertainty. However, if sufficient data is available for training, the PINN formulation can accurately infer different time-dependent fractional orders for each state of the modified SIR models.

## Model Plurality

The pluralization of epidemiological models strives for either developing a variety of compartmental models with time-dependent parameters or employing new fractional operators with different time-dependent fractional orders of the derivatives. This systematic analysis of different models provides a measure of their identifiability, predictability, and uncertainty.

A general framework in modeling the spread of infectious diseases among a fixed population is compartmental modeling, in which individuals are categorized into different epidemiological classes (compartments) according to their disease-related status. The dynamics of the virus spread through the population is governed by a nonlinear system of ODEs. The classical prototypical compartmental model is the Susceptible-Infectious-Removed (SIR) model. This simple model can be thought of as a lumped model, where the main compartments can be decomposed into various other compartments to form a variant of the SIR model to study specific state dynamics in more detail. These variations of SIR model are usually developed by appending additional compartments to the basic SIR model. The choice of the model is subjective to the purpose of the study and the available data to calibrate the model parameters, and at the present time it is done in an ad-hoc manner.

We consider different integer-order, fractional-order and time-delay models for different variations of SIR model. The integer-order models use simple derivative operators that do not account for the effect of memory in the evolution of dynamics, but instead the model parameters are treated as time-varying variables. However, many works have confirmed the power-law scaling feature of the dynamics of COVID-19 transmission^19, 20^, which indicates the notion of memory effects in the spread dynamics. In the classical SIR model, the current number of infectious individuals has exponential distribution in the infectious period, i.e., 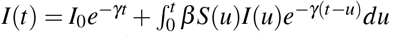. This can be interpreted as the probability for an infected subject to remain infected at time *t* is *e*^−*γt*^, where 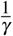 refers to the mean recovery period^21, 22^. Thus, the number of patients has exponential distribution in the mean infection period. In a general setting, we can extend the SIR model into 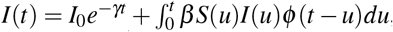, where *ϕ* is a probability function (non increasing in *t, ϕ* (0) = 1) and the mean infection period 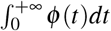^23, 24^. If we choose *ϕ* to be a step function then we arrive at a constant time-delay model, if we choose *ϕ* to be exponential function then we recover a classical SIR model^22^, and if we choose *ϕ* to be power-law waiting time distribution then we obtain a fractional SIR model^25^. Other epidemiological models characterized by partial differential equations (PDEs) can be viewed as variations of the standard ODE models that include more variables such as spatial effects, health status, and age^26–28^. While they may provide a more realistic picture of disease via spatial diffusion and higher lever parameterization, the PDE models lack structural identifiability. More importantly, the spatial data incompleteness and heterogeneity become a major issue for such models, and makes them impractical at this juncture.

In this paper, we consider a total of nine different ODE models. In a general setting, if we let **U**(*t*) be the vector of all epidemiological classes considered in a model, then the coupled system of differential equation governing the dynamics of that model can be written as

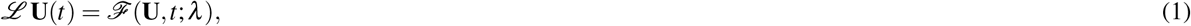

where *ℒ* is either an integer-order or a fractional-order temporal differential operator, *ℱ* is a nonlinear operator and *λ* is the set of known/unknown model parameters. The epidemiological classes in the models include the number of individuals in susceptible (**S**), exposed (**E**), pre-symptomatic (**P**), quarantined (**Q**), infectious (**I**), asymptomatic (**J**), hospitalized (**H**), death (**D**), and recovered/removed (**R**) compartments. We consider the auxiliary cumulative compartments (**I**^*c*^ and **H**^*c*^) to help fitting the available data. The three crucial time-dependent parameters are the community transmission rate (*β*_*I*_(*t*)), proportion of disease related death from the **H** class (*q*(*t*)), and proportion of hospitalized individuals (*p*(*t*)), as explained in^17^.

Seven of the nine models are shown in Fig. 1, while other two models are discussed in Supplementary Section 3. The first row in Fig. 1 shows the three integer-order variations of the classic SIR model and the transition graph between the epidemiological classes in these models. The second row in Fig. 1 shows the three fractional models with different fixed/time-variable fractional-orders. In particular, we use the Caputo fractional derivative^29^ of variable order *κ*(*t*) ∈ (0, 1) given as

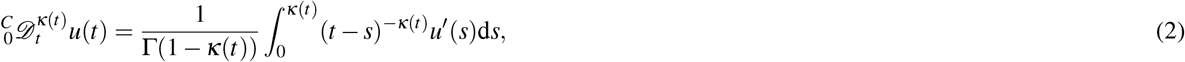

where the variable order *κ*(*t*) allows us to adjust the effect of non-locality as the dynamics evolve. A similar definition holds if the order is constant. The third row in Fig. 1 shows the time-delay model and different epidemiological classes. The detailed definition of corresponding differential equations for all of these models and the complete list of all parameters are provided in Supplementary Section 3.

**Figure 1.**
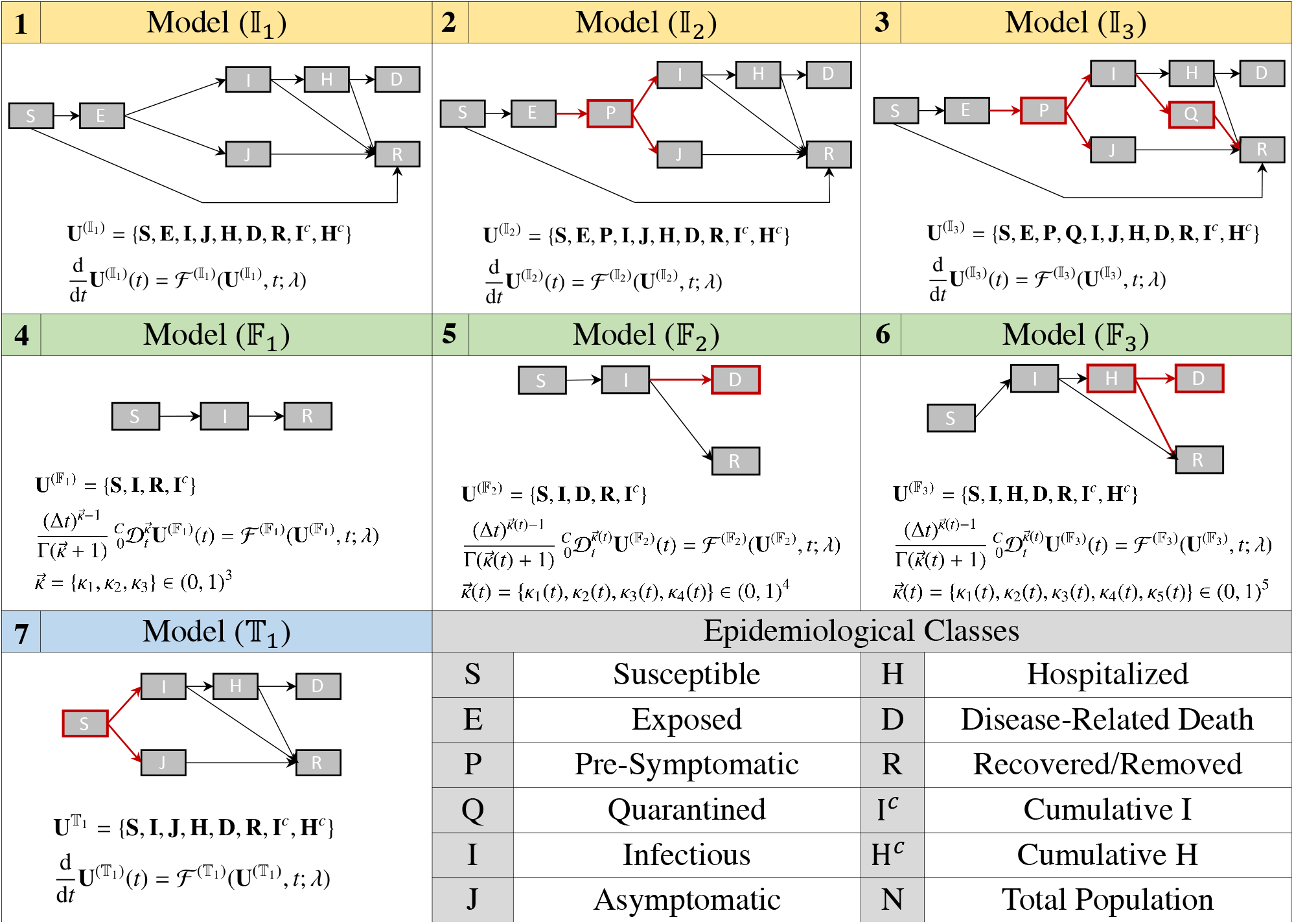
A plurality of epidemiological models. Shown are both sketches of compartments as well as their mathematical definitions. First row: Integer order models 𝕀_1_, 𝕀_2_, and 𝕀_3_. Second row: Fractional-order models 𝔽_1_, 𝔽_2_, and 𝔽_3_. Third row: Time-Delay model 𝕋_1_ and epidemiological classes. Denoted by red are the new compartments as we move from simpler to a more complex model. Details of mathematical definitions are given in the Supplementary Section 3.

## Results

We carry out our analysis based on different datasets for COVID-19 from New York City (NYC)^30, 31^, Michigan state (MI)^32, 33^, Rhode Island state (RI)^34^, and Italy^35^. In each case, the datasets are pre-processed by applying a moving averaging window of seven days to smooth the weekday-weekend fluctuations in the reporting of the outbreak. Since not all of the epidemiological states are tractable in practice, the reported data of the individuals is restricted to only a few compartments. Each dataset reports different epidemiological classes, leading to different formulation of PINNs (detailed explanation of different datasets for the PINN formulation is given in Supplementary Section 5). For example, NYC data reports the cumulative infectious **I**^*c*^, hospitalized **H**^*c*^, and death **D**^*c*^ cases. Instead, MI data reports the current values of hospitalized individuals **H**, while RI data reports both the current **H** and the cumulative hospitalized individuals **H**^*c*^. The Italy dataset reports the current **I, R**, and **D**. Given the cumulative values, we can obtain the daily increases (new) values for infectious, hospitalized, and deaths cases by **I**^*new*^(*t*) = **I**^*c*^(*t*) − **I**^*c*^(*t* − 1), **H**^*new*^(*t*) = **H**^*c*^(*t*) − **H**^*c*^(*t* − 1), and **D**^*new*^(*t*) = **D**^*c*^(*t*) − **D**^*c*^(*t* − 1).

In the following, we discuss different aspects of our formulation and results based on NYC and Italy datasets. We provide the results based on other datasets in Supplementary Section 4. In each case, we use the available datasets on observed epidemiological classes as training data for the PINN formulation for integer-order models 𝕀_1_, 𝕀_2_, 𝕀_3_, time-delay model 𝕋_1_, and fractional models 𝔽_1_, 𝔽_2_ and 𝔽_3_. We briefly discuss our methodology and PINN formulations in Methods section below. The details on the definition of the loss function for each case and the corresponding model, the structure of the neural network in the PINN formulation, and tuning the hyperparameters of the neural network are presented in Supplementary Section 5. We consider the effect of vaccination in models 𝕀_1_, 𝕀_2_, 𝕀_3_, and 𝕋_1_ by adding the extra connection from **S** to **R** with the rate 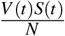. Here, *V* (*t*) denotes the number of effective vaccinated individuals at time *t*, which is calculated by *V* (*t*) = 0.52(*D*_1_(*t*) − *D*_2_(*t*)) + 0.95*D*_2_(*t*)^17^. The terms *D*_1_ and *D*_2_ denote the number of individuals that have received the first and second doses of vaccine, respectively. The first dose of COVID-19 vaccine in the US was administered on December 14, 2020. Accounting for two weeks delay in building immunity after the first dose of vaccination, we assume that the number of effective vaccinated individuals to be zero before January 1, 2021.

### Simultaneous inference of time-dependent parameters and unobserved dynamics

The PINN formulation encodes the mathematical model into the network and forms the loss function comprised of two parts. The first part is the mismatch between the network output and the available data while the second part is the ODE residuals. By minimizing the loss function, we optimize the network parameters to learn the data and simultaneously infer the unobserved dynamics by satisfying the ODEs. The success of PINN in this setting lies in its flexibility in allowing the model parameters to be time-dependent functions represented by separate neural networks. We note that in the fractional models, we infer the time-dependent fractional orders as we fix the parameters and let the fractional operators change in time.

We train PINNs for integer-order models 𝕀_1_, 𝕀_2_, 𝕀_3_, and time-delay model 𝕋_1_ based on the NYC data^31^. In Fig. 2 Panel **A**, we show the fitting of PINN formulations for the four models to the available data. The trained network can accurately fit the different fluctuations in the data. In Panel **B** and **C**, we show the inferred time-dependent parameters (*β*_*I*_(*t*), *p*(*t*), *q*(*t*)) and the inferred unobserved compartments for the four models, respectively. The plots demonstrate an almost similar trend throughout the spread of the virus. Some of the models have additional compartments, e.g., the **Q** compartment in model 𝕀_3_.

**Figure 2.**
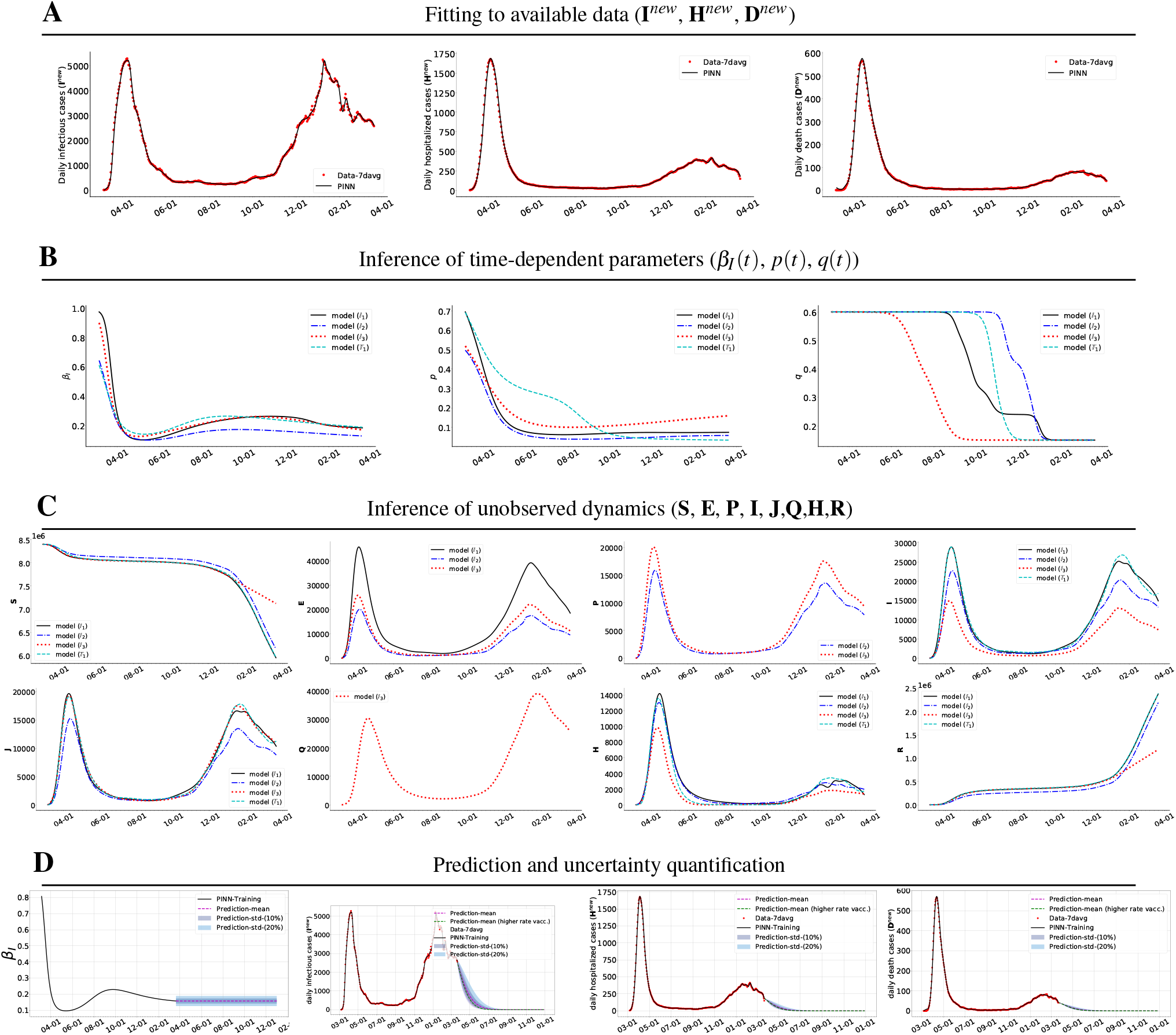
PINNs inference using the integer-order models 𝕀_1_, 𝕀_2_, 𝕀_3_ and time-delay model 𝕋_1_ for NYC. **A**: Accurate fitting to the available data of daily infectious, hospitalized, and death cases. **B**: Inference of time-dependent parameters (*β*_*I*_(*t*), *R*_*c*_(*t*), *p*(*t*), *q*(*t*)). **C**: Inference of unobserved dynamics. **D**: Prediction and uncertainty quantification of daily infectious, hospitalized, and death cases for a four-month time window using the model 𝕀_2_. Here the inferred delay is *d* = 3.10 for the time-delay model 𝕋_1_.

### Forecasting with uncertainty

The inferred model from PINNs yields a predictive model that we use to forecast future dynamics by considering different control measures for each model. In particular, we can predict the evolution of dynamics using the integer-order models 𝕀_1_, 𝕀_2_, 𝕀_3_, and the time-delay model 𝕋_1_. Panel **D** in Fig. 2 shows the prediction of new infectious, hospitalized, and death cases for NYC data based on model 𝕀_2_. In the prediction window, we fix the parameters *β*_*I*_(*t*), *p*(*t*), and *q*(*t*) at their final values from the training window. However, we postulate that the effect of several different control measures in the community transmission rate *β*_*I*_(*t*) can be captured by adding an uncertainty bound of 10% and 20% to its mean value. By using standard forward propagation techniques, i.e., Monte Carlo^36^ and probabilistic collocation methods^37^, we propagate the uncertainty into the prediction window. We also consider different vaccination scenarios by considering different vaccination rates in calculating the effective vaccination per day (*V* (*t*)) in the prediction window by

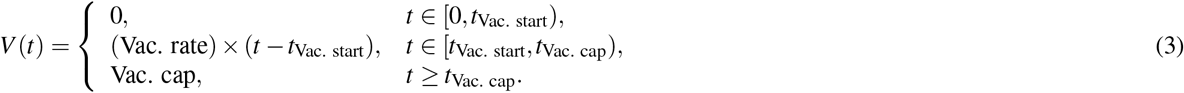

In particular, for the NYC data set^30^, we set *t*_Vac. start_ = 300 and Vac. cap = 30000. We consider two vaccination rate and adjust *t*_Vac. cap_, e.g., Vac.rate = 600 and *t*_Vac. cap_ = 350.

### Fractional Models

The structure of PINNs for fractional models is usually more complex compared to the other models. This is mainly because of the fractional operators, for which the automatic differentiation technology is not applicable and, hence, we need to resort to numerical discretization, such as the L1 scheme^38,39^. This formulation, namely fractional PINNs (fPINNs), was first developed in^40^ and we extend it to apply to our problem. The other source of the complexity of PINN structure for fractional models is the time-dependent fractional orders. The PINN formulation uses a separate neural network to represent each fractional order *κ*_*i*_(*t*). In particular, Figure 3 shows the fPINNs schematic for the fractional model 𝔽_3_. We use a (relatively large) neural network to represent the states 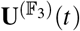 (the green shaded box) and five separate neural networks to represent the fractional orders *κ*_*i*_(*t*), *i* = 1, …, 5 (the red shaded boxes). We discuss our methodology in the Methods section below and provide the formulation in Supplementary Section 5. The PINN formulation makes the inverse problem more efficient as it uses the network to represent the time-dependent parameters and integrates data and differential equations through the loss function. Therefore, it provides a powerful computational tool to perform inference in fractional models.

**Figure 3.**
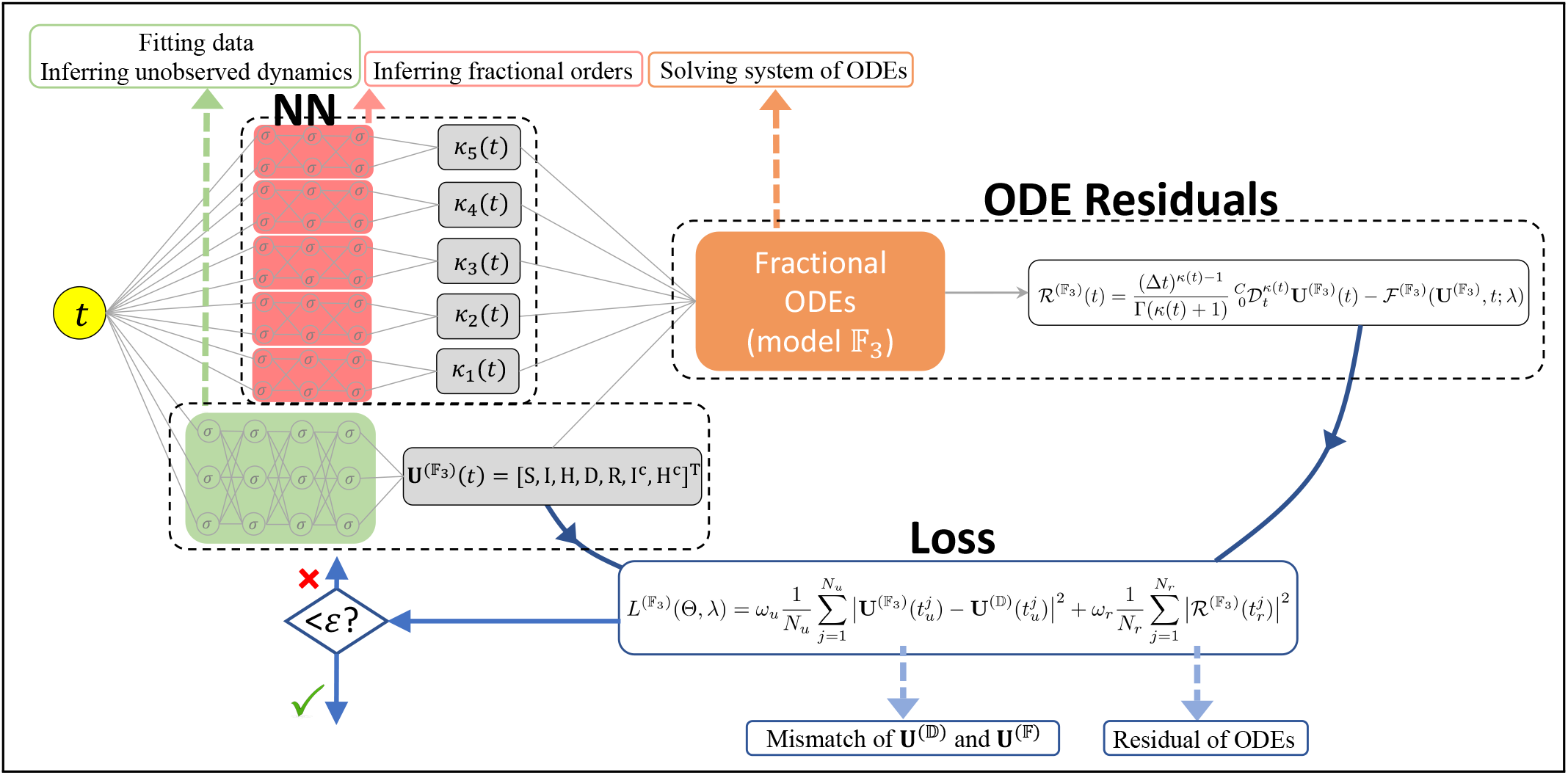
Schematic of the physics-informed neural network for the fractional model 𝔽_3_. **NN**: denotes six different neural networks. The green shaded NN represents the state 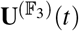 to fit the available data **U**^(𝔻)^(*t*) and infer the unobserved dynamics. The red shaded NNs represent different fractional orders *κ*_*i*_(*t*), *i* = 1, …, 5 to infer their time-dependence. **ODE Residuals**: computing the residual of fractional model 𝔽_3_. **Loss**: is comprised of two parts; the mismatch between available data and NN output and the ODE residual. By minimizing the loss function 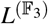, the NNs simultaneously fit the data *and* infer the unobserved dynamics and fractional orders by satisfying the system of ODEs. See Fig. 5 in Supplementary Section 5 for similar PINNs for integer-order models.

From the available data for NYC, we only have the new cases infectious **I**^*new*^, hospitalized **H**^*new*^, and deaths **D**^*new*^. This is a limited amount of data for the fractional model 𝔽_3_ with five unobserved compartments and five time-dependent parameters so it leads to an identifiability problem for this model. We present this issue in Fig. 4 via numerical experiments. We consider the fractional model 𝔽_3_ and its integer-order counterpart (by letting *κ*_*i*_(*t*) = 1, *i* = 1, …, 5 and representing *β*_*I*_(*t*) by a separate neural network) for comparison. We plot the results of fractional model 𝔽_3_ against the results of the integer-order counterpart in panels **A**-**C** by employing different amounts of data. In each panel in the top row, black color shows the results of model 𝔽_3_ while red color shows the results of the corresponding integer-order model.

**Figure 4.**
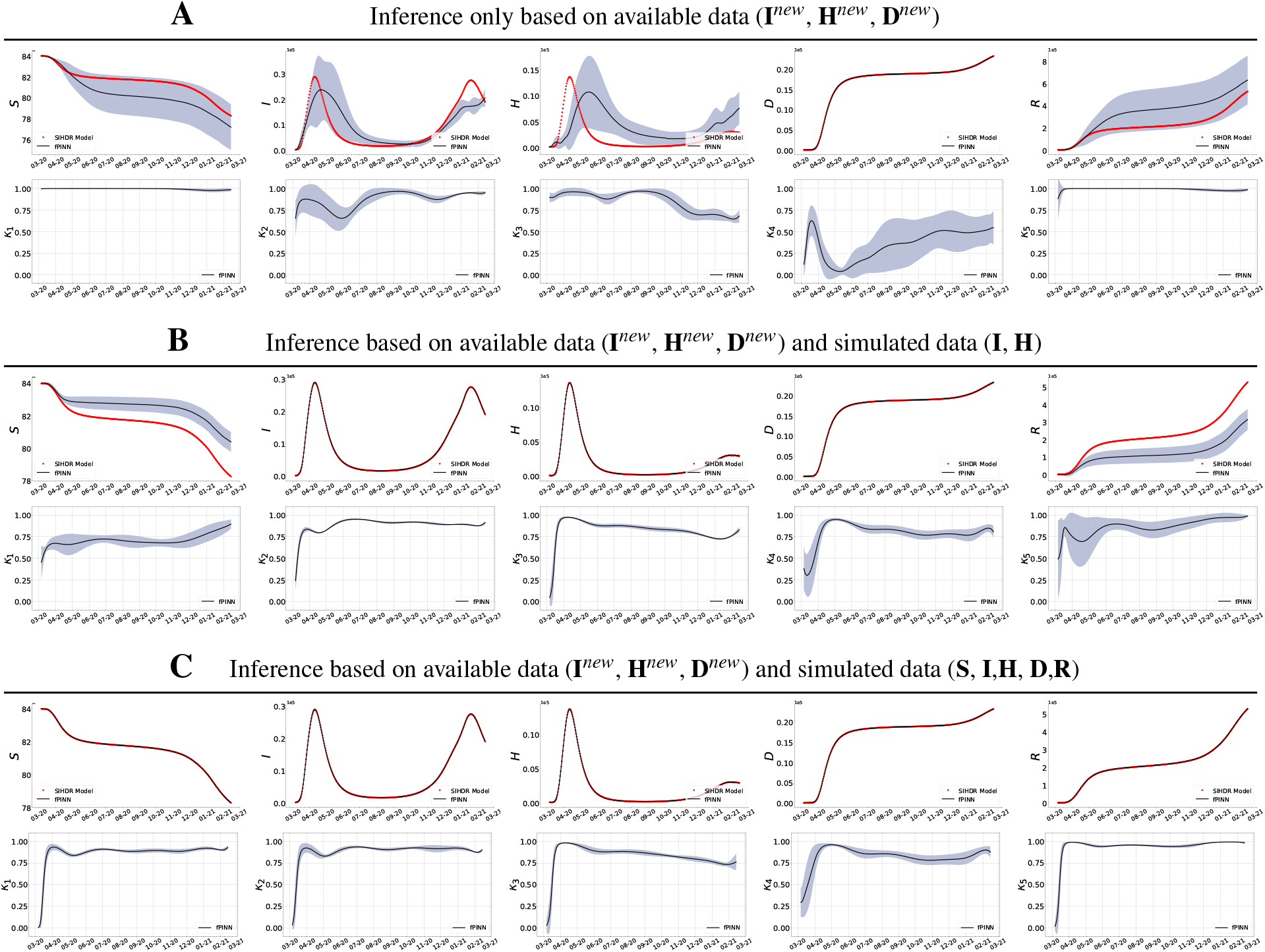
Identifiability study for PINNs for the fractional order model 𝔽_3_. Instead of time-dependent parameters, here we aim to infer different time-dependent fractional operators for *κ*_*i*_(*t*), *i* = 1, …, 5. In **A**-**C**, in the first row we plot the inferred unobserved dynamics and in the second row the inferred corresponding time-dependent fractional orders for different available data for NYC. **A**: We employ only the available data on *I*^*new*^, *H*^*new*^, and *D*^*new*^ for NYC. **B**: In addition to **A**, we also assume that we have additional simulated data on I and H. **C**: We have additional simulated data on S, I, H, D, and R. The uncertainty bands shown reflect the robustness of each case.

In Fig. 4 panel **A**, we only use the available data (**I**^*new*^, **H**^*new*^, **D**^*new*^) to train the network. We can clearly observe that the inferred dynamics and the corresponding fractional order have relatively large uncertainty bounds and are different from the results of the integer-order model. We note, however, that the **D** compartment has the smallest uncertainty bound. This is because the **D** compartment does not have any outflow and hence **D** = **D**^*c*^. It is counter-intuitive, however, that its corresponding fractional order has the largest uncertainty bounds. This is due to the coupling of dynamics of the **D** compartment with other **H** and **R** compartments that contain large uncertainty bounds. In panel **B**, in addition to the data in panel **A**, we use the simulated data of **I** and **H** compartments. These values are not available in practice for the NYC data, and hence we obtain them from the corresponding integer-order model and use them to investigate the identifiability of the fractional model 𝔽_3_. We can see that the inference can be improved by including the additional simulated data. In panel **C**, we show that if we add extra data we can further reduce the uncertainty bounds in the inferred time-dependent parameters. It is interesting to observe in panel **C** that a fractional model with time-dependent fractional order can resemble the dynamics of its integer-order counterparts with time-dependent parameters. This example supports the two alternative paradigms of modeling philosophy, discussed earlier in the introduction section.

We note that although the fractional models are capable of accounting for memory effects via non-local operators and their application has been motivated in the literature to a great extent, the crucial less-studied element is to develop a robust computational framework to implement the inverse problem of inferring the time-dependent fractional orders. For example,^19^ suggests the use of time-dependent variables fractional orders, however, in their implementation the fractional orders are parameterized with a few constant parameters as piece-wise constant functions to render the inverse problem feasible in their formulation. The under-parametrization of fractional orders may lead to inaccurate results. More importantly, when different fractional orders are considered, the total population is not conserved automatically by the system of ODEs and should be enforced in the simulation; see the Discussion section for more details. We show here that our formulation has the capacity to efficiently extend the parameterization of time-dependent fractional orders via neural networks, and we implement an efficient computational framework to infer them from data. For comparison, we consider the identifiability of fractional model 𝔽_2_ based on the Italy data. This is a practical example of the case discussed above in Fig. 4 panel **C**. The Italy data reports the current values **I, R**, and **D** (and **S** = *N*— **I**−**D**−**R**), which are the four compartments in the fractional model F_2_ with time-dependent fractional orders. Since we have access to a large amount of data in this case, we do not fix the infection rate parameter (*r*(*t*), see Tables 1 and 3 in Supplementary Section 3) and let it be represented by a separate neural-network. Figure 5 left column shows the accurate fitting to the available data and the right column shows the inferred time-varying parameters *κ*_*i*_(*t*), *i* = 1, 2, 3, 4, and *r*(*t*). These results demonstrate that even in the case of access to data for all compartments, the inference of fractional orders is not very robust and still exhibits some uncertainties, which is a reflection of lack of identifiability of the model. The choice of piece-wise constant parameters in the work of^19^ simplifies the problem but leads to erroneous results.

**Figure 5.**
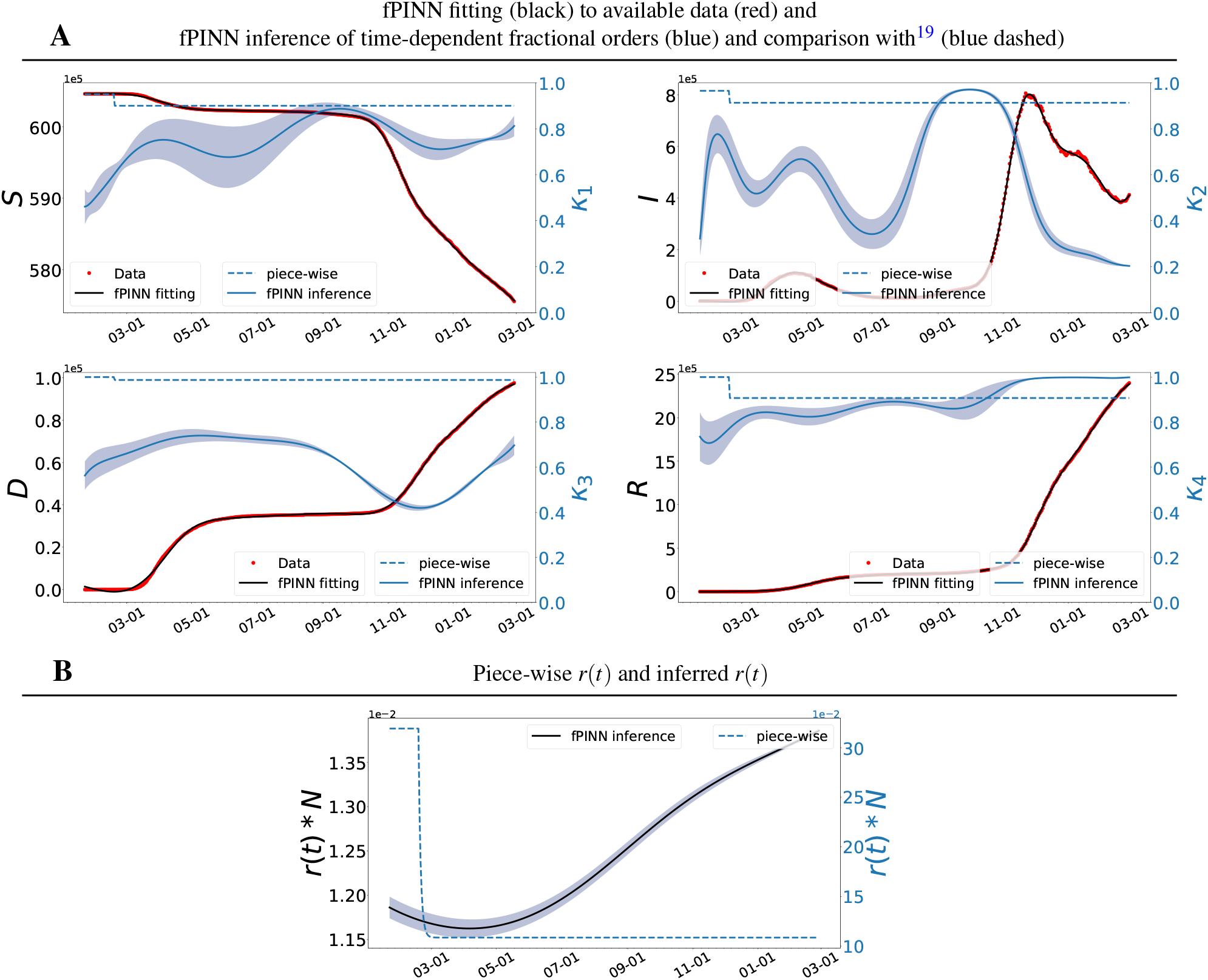
PINNs results of the fractional order model 𝔽_2_ based on Italy data. In **A**: we show accurate fitting (black) to the available data **S, I, R**, and **D** (red). In each sub-figure, we also show fPINN inference of the corresponding time-dependent fractional order (blue) and compare it with the piece-wise fractional orders (dashed blue). In **B**: we compare the piece-wise parameter *r*(*t*) and the fPINN inference of time-dependent parameter *r*(*t*). Here the piece-wise fractional orders and parameters are given by^19^. The other parameters of the model are fixed as *a* = 0.0215 and *b* = 0.0048^19^ in the simulation.

## Method

PINNs were first introduced in^16^ and since then PINNs have been successfully applied in solving forward and inverse problems in many practical applications^41^–48. The analysis and convergence study of PINNs have been carried out in^49^. Figure 3 shows the schematic of PINNs, which is almost the same for all of the models that we consider in this paper. The depicted structure in Fig. 3 is specifically related to the fractional order models 𝔽_3_ with five time-dependent parameters.

In the PINN formulation, we use separate deep neural networks with input *t* to represent the states **U**(*t*) and (time-dependent) parameters. Each network is parameterized by a set of parameter Θ as weights and biases of the network. Thus, we let **U**(*t*) ≈ **U**_*NN*_(*t*; Θ_*U*_) (the green shaded box in Fig. 3). For integer-order models 𝕀_1_, 𝕀_2_, 𝕀_3_, time-delay model 𝕋_1_, we let 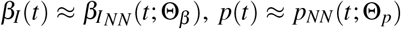, and *q*(*t*) ≈ *q*_*NN*_(*t*, Θ_*q*_). For fractional order models 𝔽_1_, 𝔽_2_, 𝔽_3_, we let 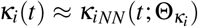 for *i* = 1, …, 5 (the red shaded boxes in Fig. 3). By using (1), we define the residual of equation as *ℛ*_*NN*_(*t*) = *ℒ* **U**_*NN*_(*t*) − *ℱ*(**U**_*NN*_, *t*; *λ*) (the orange shaded box in Fig. 3), where *ℒ* is the integer-order, time-delay, or fractional-order differential operator. This residual is the measure of the approximation **U**_*NN*_ satisfying the ODEs, and ideally the exact solution is recovered when the residual is identically zero. We define two finite sets of training points 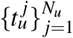 and residual points 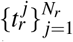.The training points are the points where data is available and the residual points are the points where the residual *ℛ*_*NN*_(*t*) is satisfied and they are freely available all over the computational domain. Therefore, we define the loss function of PINN as

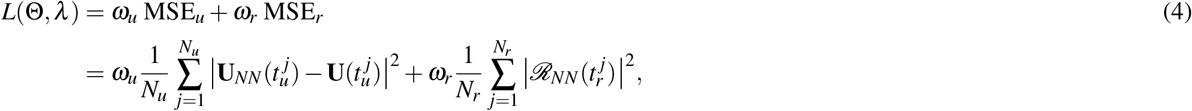

where MSE stands for mean squared error. The loss function of PINNs is comprised of two terms: MSE_*u*_ and MSE_*r*_, which are defined by the last part of the above equation. The parameters {*ω*_*u*_, *ω*_*r*_} denote the weight coefficients in the loss function that can balance the optimization effort between learning the data and satisfying the ODEs. They may be user-specified or tuned manually or automatically, e.g., in practice based on the numerical experiment in each problem^50^.

In all cases, the derivatives of the network with respect to the input *t* and network parameters Θ are computed by applying the chain rule for differentiating compositions of functions using the automatic differentiation^51^. In particular, we use Tensorflow programming^52^ for automatic differentiation and deep learning computations. The detailed derivation of loss function for different models and different datasets are provided in Supplementary Section 5.

## Discussion

The surge of COVID-19 disease prompted many researchers around the world to analyze the dynamics of disease transmission by employing different statistical and epidemiological models, providing the opportunity to further revise and modify to some extent the underlying assumptions, limitations, robustness, and the associated uncertainties of such models. While studying the behavior of virus in a pandemic presents a broader opportunity to interrogate how to manage pathogens in future events^4^, critical investigation of different epidemiological models can also shed light on their advantages, limitations, and sensitivity to small changes^53^. Appropriate mathematical models can be used to estimate parameters of pathogen spread, explore possible future scenarios for control measures, evaluate retrospectively the efficacy of specific interventions, and identify prospective strategies^54^. However, the main problem with these lumped epidemiological models is the lack of uniqueness of parameters for integer-order models and fractional operators for fractional-order models. At the present time, there are no rigorous a priori identifiability analysis for models with time-dependent parameters or time-dependent fractional orders. Hence, in the present work we resorted to systematic numerical experimentation to provide a posteriori some measures of identifiability using uncertainty quantification. We have learned some useful lessons by fitting the epidemic data using nine different epidemiological models (five integer-order, one time-delay, and three fractional-order models; see Supplementary Section 3). In the following, we summarize some observations related to our findings.

### Sources of uncertainty

The novelty of coronavirus naturally leads to many uncertainties^1^. This is due to many unknown factors including biological features of transmission, different mutation of virus and pathogen behavior and concentrations. The most systematic source of uncertainty that can adversely affect almost all mathematical models output is that the exact number of infected individuals is unknown. Always, the onset of outbreak proceeds the start of reporting data, causing a lack of initial data at the beginning of the outbreak and inaccurate parameter estimation in all models during that initial period, especially those with memory effects. For example, if we use the parameters estimated in^55^ for fitting data up to present time, the results are totally erroneous. The reported data contains the intermittent artifacts due to weekdays/weekends effects; see Fig. 6 panel **A**. Hence, these inherent fluctuations of collected data is another key factor in the uncertainty of the inverse problem. The reporting practices of COVID-19 case data are not consistent among different populations. For example, what is reported in NYC is different than in RI despite the geographic proximity. The same model may perform differently if applied to datasets from those two sources. Therefore, because of the multitude of uncertainties, quantifying the parametric input uncertainty is not sufficient^5^. Consequently, the model selection becomes the major issue, in our opinion, not only on how to best fit the data but also on how robust the model is given these uncertainties.

**Figure 6.**
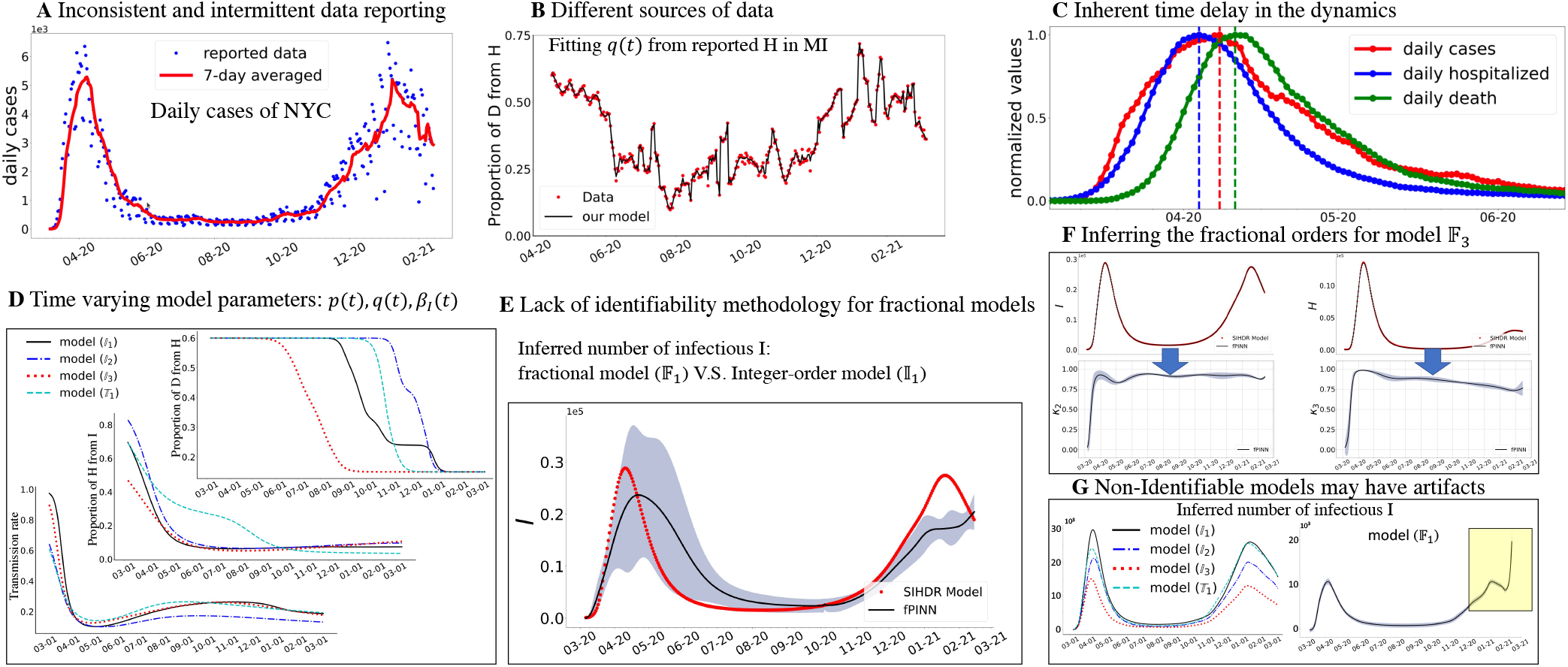
Main challenges for epidemiological models highlighted in A-G. **A**: Number of daily infectious cases (**I**^*new*^) for NYC, with blue representing the raw reported data and red representing the seven-day average. **B**: Additional available data of current hospitalized individuals for MI to fit the parameter *q*(*t*) expressing proportion of disease related deaths from the **H** class which is only available in some states. **C**: Inherent time-delay in the only available data for NYC. **D**: Inferred time-varying parameters (*β*_*I*_(*t*), *p*(*t*), *q*(*t*)) of integer-order and delay models 𝕀_1_, 𝕀_2_, 𝕀_3_, and 𝕋_1_ based on NYC data. **E**: While there are a priori identifiability analyses for integer-order models, there is no identifiability methodology for fractional models. Shown is a comparison of inferred dynamics of models 𝔽_1_ and 𝕀_1_ for NYC data. The shaded region is the uncertainty of the fractional model. **F**: Inferred different time-dependent fractional orders of model 𝔽_3_ using simulated data based on the integer-order model for NYC. **G**: On the left we plot the inferred dynamics from integer-order and time-delay models that show differences but consistent trends. On the right, we show the inferred dynamics of model 𝔽_1_ that shows an erroneous trend due to lack of indentifiability (window in yellow). The SIHDR model in Panel **E** is defined as model 𝕀_5_ in Supplementary Section 3.4.

### Use all the data

The compartmental models describe the outbreak by considering the current number of individuals in each epidemiological class. However, the reported data usually contain the daily counts (new cases) or the cumulative number of individuals. Therefore, in order to fit the available data, additional auxiliary equations should be added to these models. If the current values of the epidemiological classes are reported (such as hospitalization in MI data^32^), then some of the model parameters can be directly obtained (the proportion of disease-related deaths from the hospitalization, i.e., *q*(*t*)). This is shown in Fig. 6 panel **B**.

### Inherent time-delay

The time history of reported data for different epidemiological classes shows a time-delay in the transition of individuals from one compartment to another, especially during the outbreak surge; see Fig. 6 panel **C**. This implies that the mathematical model should have the capacity to account for the delayed dynamics, as we did with our time-delay integer-order model, where the delay constant was determined directly by the data. Moreover, as the disease symptoms, concentration and behavior of pathogen are changing throughout the course of the disease^56,57^ the models should accommodate time-dependent parameters. In our formulation, the integer-order models account for this time-variability; see Fig. 6 panel **D**. Additionally, the fractional models include different memory effects for each epidemiological class by considering different time-dependent fractional orders; see Fig. 6 panel **F**.

### Identifiability for fractional models

The identifiability of integer-order models with piece-wise constant parameters has been systematically investigated in^17^. However, such analysis is not available for the fractional models, which may exhibit higher uncertainty; see Fig. 6 panel **E**. Here, we investigated the robustness of fractional-order models and we have concluded that due to their fragility, should any fractional model is considered, identifiability must be investigated at least numerically. Even if fitted accurately to the available data, a non-identifiable model may still show some serious artifacts that are not intuitively consistent; see Fig. 6 panel **G**.

### Conservation of total population

In most integer-order models, the conservation of the total population is satisfied automatically by the system of ODEs. This is not the case for the fractional models when different fractional orders are considered. We have observed that in many published studies using the fractional modeling, this fact has been overlooked and the population conservation has not been included in the system of equations. This can result in an erroneous output of the model, especially since compartmental models require this assumption.

### Dimensional consistency in fractional models

The dynamics of each compartment in the fractional epidemiological model can be obtained by 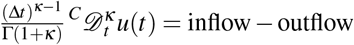, where the time scale Δ*t* denotes a characteristic time of observation which amounts to a built-in scale effect. The scale effect goes away as *κ* → 1, Δ*t*^*κ*−1^ → Δ*t*^0^ → 1, and thus the equation recovers the classical continuity equation. This scaling factor is also useful in practice as it ensures the dimensional consistency of the units in the fractional model (see Supplementary Section 3.3 for more detail).

### Long term prediction

The purely statistical approaches are generally not well suited for long-term predictions of epidemiological dynamics; see e.g. Fig. 1 in Supplementary Section 2. They do not have the ability to account for how the transmission occurs and thus most of the forecasting models limit their projections to one week or a few weeks ahead^58^. The mechanistic models, if their parameters are inferred correctly from data, are often helpful to quantify different sources of uncertainty and examine the implications of various assumptions and control measures about a highly nonlinear process that is hard to predict using only intuition or statistical models. However, they are constrained by their limitations, what we assume, and what we do not know. For example, the proportion of hospitalized individuals, *p*(*t*), may reach an asymptotic value after some time, but the community transmission rate, *β*_*I*_(*t*), is highly influenced by control measures, mobility, mutants variations, etc, and hence we specify it as a random variable for future predictions, see Fig. 2 panel **D**.

In summary, given the aforementioned observations, we recommend the use of multiple models tailored to specific region and specific data availability, and quantify both parametric uncertainty as well as model uncertainty.

## Data Availability

All sources of data are referenced in the manuscript.

## Data availability

All different data sets used in this paper are publicly available and are downloaded/obtained from^31,32,34,35^.

## Code availability

The version of PINN-COVID that was used to generate our results is publicly available at a GitHub branch.

## Acknowledgements

MC acknowledges support by the China Scholarship Council (No. 201906890040). GEK and GL acknowledge support by the MURI/ARO, W911NF-15-1-0562.

## Author contributions

GEK, GL and EK designed the study and supervised the project. EK and MC developed the integer- and fractional-models and ran the simulations. XZ developed the time-delay model and ran the simulations. All authors analyzed the results and contributed to the writing of the paper.

## Additional information

**Supplementary information** The online version contains supplementary material.

