## Supplementary Information for "Identifiability and predictability of integer- and fractional-order epidemiological models using physics-informed neural networks"

### 1 Introduction

The Supplementary Information contains additional results that provide more information about the motivation of the work, definition and details of the nine epidemiological models considered in this study, detailed discussion on different sets of results, and detailed formulation of physics-informed neural networks (PINNs) for integer- and fractional-order models.

### 2 Model uncertainty

Quantifying parametric input uncertainty is not sufficient and the effect of model structure must be properly studied[1]. Figure 1 clearly shows the uncertainty associated with several different models in analysing and predicting the dynamics of this complex system. The figure is obtained from the COVID-19 Forecast Hub[2], which is a public online server that serves as a central repository of forecasts and predictions from over 50 international research groups.

### 3 Details of different epidemiological models

In a general setting, if we let $\mathbf{U}(t)$ be the vector of all epidemiological classes considered in a model, then the coupled system of differential equation governing the dynamics of that model can be written as

$$\mathscr{L}\,\mathbf{U}(t) = \mathscr{F}(\mathbf{U},t;\lambda), \tag{1}$$

where $\mathscr{L}$ is an integer-order or fractional-order temporal differential operator, $\mathscr{F}$ is a nonlinear operator and $\lambda$ is the set of known/unknown model parameters. Here, we define and provide details on nine different models with various compartments. The complete list of all parameters used in the models is given in Table 1.

#### 3.1 Integer-Order Models

We consider three integer-order variations of the classic SIR model by introducing more epidemiological classes. The governing equations are given in Table 2.

• Model $\mathbb{I}_1$: Integer-order SEIJDHR. We divide the total population into seven different compartments and let $\mathbf{U}^{(\mathbb{I}_1)}(t) = \{\mathbf{S},\mathbf{E},\mathbf{I},\mathbf{J},\mathbf{H},\mathbf{D},\mathbf{R},\mathbf{I}^c,\mathbf{H}^c\}$. Therefore, the governing equation of the dynamics can be written as $\frac{\mathrm{d}}{\mathrm{d}t}\mathbf{U}^{(\mathbb{I}_1)}(t) = \mathscr{F}^{(\mathbb{I}_1)}(\mathbf{U}^{(\mathbb{I}_1)},t;\lambda)$; see Table 2 first column. The effective reproduction number for model $\mathbb{I}_1$ is given by $\mathscr{R}_c = \beta_I\left(\dfrac{(1-\delta)\varepsilon}{\gamma_a} + \dfrac{\delta}{\gamma}\right)$.

• Model $\mathbb{I}_2$: Integer-order SEPIJDHR. We include the pre-symptomatic individuals by adding an extra compartment $\mathbf{P}$ in model $\mathbb{I}_1$. Therefore, we let $\mathbf{U}^{(\mathbb{I}_2)}(t) = \{\mathbf{S},\mathbf{E},\mathbf{P},\mathbf{I},\mathbf{J},\mathbf{H},\mathbf{D},\mathbf{R},\mathbf{I}^c,\mathbf{H}^c\}$ and thus the governing equation of the dynamics can be written

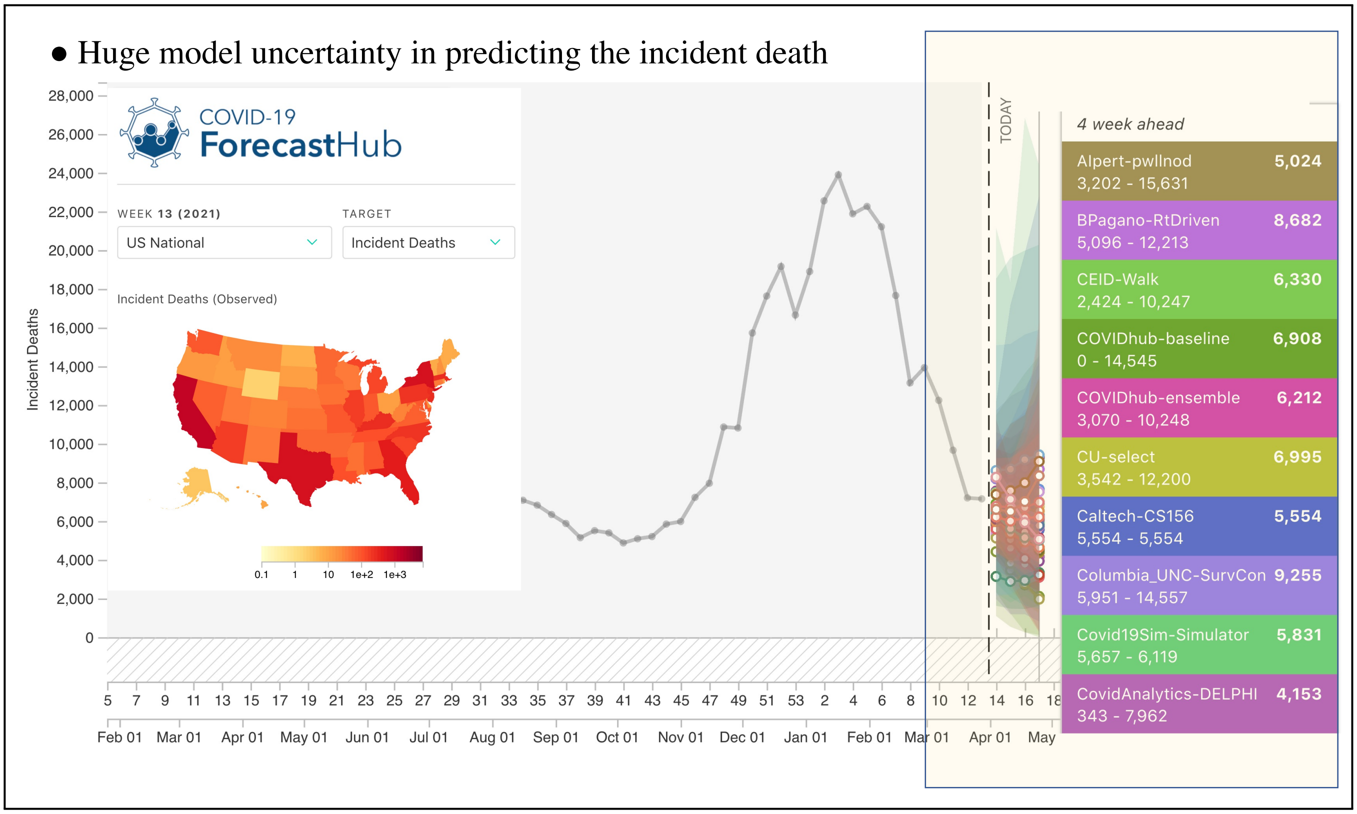


**Figure 1. Prediction uncertainty associated with different models.** The figure is obtained from the COVID-19 Forecast Hub[2], which is a public online server that serves as a central repository of forecasts and predictions from over 50 international research groups.

as $\frac{\mathrm{d}}{\mathrm{d}t}\mathbf{U}^{(\mathbb{I}_2)}(t) = \mathscr{F}^{(\mathbb{I}_2)}(\mathbf{U}^{(\mathbb{I}_2)}, t; \lambda)$; see Table 2 second column. The effective reproduction number for model $\mathbb{I}_2$ is given by $\mathscr{R}_c = \beta_I \left( \dfrac{(1-\delta)\varepsilon}{\gamma_a} + \dfrac{\delta}{\gamma} + \dfrac{\chi}{\alpha_2} \right)$.

• Model $\mathbb{I}_3$: Integer-order SEPIJDHQR. We include the pre-symptomatic and quarantined individuals by adding extra compartments $\mathbf{P}$ and $\mathbf{Q}$ in model $\mathbb{I}_1$. Therefore, we let $\mathbf{U}^{(\mathbb{I}_3)}(t) = \{\mathbf{S}, \mathbf{E}, \mathbf{P}, \mathbf{Q}, \mathbf{I}, \mathbf{J}, \mathbf{H}, \mathbf{D}, \mathbf{R}, \mathbf{I}^c, \mathbf{H}^c\}$ and thus the governing equation of the dynamics can be written as $\frac{\mathrm{d}}{\mathrm{d}t}\mathbf{U}^{(\mathbb{I}_3)}(t) = \mathscr{F}^{(\mathbb{I}_3)}(\mathbf{U}^{(\mathbb{I}_3)}, t; \lambda)$; see Table 2 third column. The effective reproduction number is given by $\mathscr{R}_c = \beta_I \left( \frac{(1-\delta)\varepsilon}{\gamma_a} + \frac{\delta}{d_I} + \frac{\varepsilon}{\alpha_2} \right)$.

#### 3.2 Time-Delay Model

• Model $\mathbb{T}_1$: Time-delay SIJHDR. In this model, we simplify model $\mathbb{I}_1$ by removing the $\mathbf{E}$ compartment and introducing a delay in the dynamics of $\mathbf{S}$ compartment. We divide the total population into six different compartments and let $\mathbf{U}^{\mathbb{T}_1}(t) = \{\mathbf{S}, \mathbf{I}, \mathbf{J}, \mathbf{H}, \mathbf{D}, \mathbf{R}, \mathbf{I}^c, \mathbf{H}^c\}$. The governing equation of the dynamics can be written as $\frac{\mathrm{d}}{\mathrm{d}t}\mathbf{U}^{(\mathbb{T}_1)}(t) = \mathscr{F}^{(\mathbb{T}_1)}(\mathbf{U}^{(\mathbb{T}_1)}, t; \lambda)$; see Table 4. Similar to the integer-order model $\mathbb{I}_1$, the effective reproduction number for model $\mathbb{T}_1$ is given by $\mathscr{R}_c = \beta_I \left( \dfrac{(1-\delta)\varepsilon}{\gamma_a} + \dfrac{\delta}{\gamma} \right)$.

#### 3.3 Fractional-Order Models

We consider the Caputo fractional derivative[14] of (variable) order $\kappa(t) \in (0,1)$ given as

$$ {}_0^C\mathscr{D}_t^{\kappa(t)} u(t) = \frac{1}{\Gamma(1-\kappa(t))} \int_0^{\kappa(t)} (t-s)^{-\kappa(t)} u'(s) \mathrm{d}s, \tag{2} $$

where similar definition holds if the order is constant. To derive the fractional model, we first consider the fractional conservation of mass equation[15]. Let $u(t)$ denote the dynamics of a complex system defined on the half-time axis $t \in [0, \infty]$. The left-sided

**Table 1.** Parameters of Epidemiological models.

| Symbol | Definition | Value/Range | Note/Reference |
|---|---|---|---|
| $\beta_I$ | Community transmission rate | (0, 1) | Fixed/Fitted |
| $q$ | Proportion of disease related deaths from the H class | (0, 1) | Fixed/Fitted |
| $p$ | Proportion of hospitalized individuals | (0, 1) | Fixed/Fitted |
| $r$ | infection rate | (0, 1) | Fitted |
| $a$ | recovery rate | (0, 1) | Fixed |
| $b$ | death rate | (0, 1) | Fixed |
| $d$ | number of delay days in time-delay model | [0, 10) | Fitted |
| $\varepsilon$ | Infectivity ratio of mildly infected to severely infected | 0.75 | [3] |
| $\delta$ | Proportion of asymptomatic infections | 0.6 | [3,4] |
| $\chi$ | Infectivity ratio of pre symptomatic to symptomatic | 0.55 | [5] |
| $\gamma_1$ | removal rate | 0.0365 | [6] |
| $1/\alpha$ | Mean Incubation period | 5.2 days | [3,7,8] |
| $1/\alpha_1$ | Latent period | 2.9 days | [5] |
| $1/\alpha_2$ | Pre-symptomatic infectious period | 2.3 days | [5] |
| $1/\gamma_a$ | Mean infectious period for J class | 6 days | [9–11] |
| $1/\gamma$ | Mean symptomatic infectious period | 6 days | [9–11] |
| $1/\phi_R$ | mean duration in H before disease recovery | 7.5 days | [3] |
| $1/\phi_D$ | mean duration in H before disease death | 15 days | [12] |
| $1/d_I$ | Mean duration in I before quarantine | 2.9 days | [5] |
| $1/d_H$ | Mean duration in H class | 6.9 days | [3] |
| $1/d_Q$ | Mean duration in Q class | 10 days | [13] |

Caputo-Taylor series of $u(t)$ is given as

$$u(t+\Delta t) = u(t) + \frac{(\Delta t)^{\kappa}}{\Gamma(1+\kappa)}\, {}^{C}\mathscr{D}_t^{\kappa} u(t) + \frac{(\Delta t)^{2\kappa}}{\Gamma(1+2\kappa)}\, {}^{C}\mathscr{D}_t^{\kappa}\, {}^{C}\mathscr{D}_t^{\kappa} u(t) + \cdots, \tag{3}$$

where $\kappa \in \mathbb{R}^+$ and the expansion includes the sequential fractional derivative. It has been shown in[15] that if the change in mass flux follows a power-law of order $\kappa$ and if we match the order of the fractional Taylor series approximation to the exponent in the power law function, then the two-term fractional Taylor series approximation to this function is exact. By truncating the expansion after second term, we can approximate the rate of change of $u(t)$ by

$$\frac{u(t+\Delta t)-u(t)}{\Delta t} \approx \frac{(\Delta t)^{\kappa-1}}{\Gamma(1+\kappa)}\, {}^{C}\mathscr{D}_t^{\kappa} u(t), \tag{4}$$

which is equal to the net mass flux of the system. Therefore, for each compartment in the epidemiological model, we can write

$$\frac{(\Delta t)^{\kappa-1}}{\Gamma(1+\kappa)}\, {}^{C}\mathscr{D}_t^{\kappa} u(t) = \text{inflow} - \text{outflow}. \tag{5}$$

The time scale $\Delta t$ appears in (5) denotes a characteristic time of observation which amounts to a built-in scale effect. It is usually considered to be the total time of observation[16]. The scale effect goes away as $\kappa \to 1$, $\Delta t^{\kappa-1} \to \Delta t^0 \to 1$ and thus the equation recovers the classical continuity equation. This scaling factor is also useful in practice as it ensures the consistency of units' dimensions in fractional models. Here, we take $\Delta t = 7.0$ in our simulation since we pre-process the data by taking a seven-day average. We consider three fractional models. We note that the fractional operators can be of different orders for each compartment, and they can be either fixed or time-dependent parameters.

• Model $\mathbb{F}_1$: Fractional-order SIR. We use the Caputo fractional derivative of different fractional orders for each compartment in the integer-order classical SIR model. We let $\mathbf{U}^{(\mathbb{F}_1)}(t) = \{\mathbf{S}, \mathbf{I}, \mathbf{R}, \mathbf{I}^c\}$ and thus the governing equation of the dynamics takes the form $\frac{(\Delta t)^{\vec{\kappa}-1}}{\Gamma(\vec{\kappa}+1)}\, {}_0^C\mathscr{D}_t^{\vec{\kappa}} \mathbf{U}^{(\mathbb{F}_1)}(t) = \mathscr{F}^{(\mathbb{F}_1)}(\mathbf{U}^{(\mathbb{F}_1)}, t; \lambda)$; see Table 3 first column. Here, $\vec{\kappa} = (\kappa_1, \kappa_2, \kappa_3) \in (0,1)^3$ with $\kappa_1$, $\kappa_2$, and $\kappa_3$ being the fractional derivative orders for the compartments $\mathbf{S}$, $\mathbf{I}$, and $\mathbf{R}$, respectively. The effective reproduction number is given by $\mathscr{R}_c = \frac{\beta_I}{\gamma_1}$.

• Model $\mathbb{F}_2$: Fractional-order SIDR. We include the additional compartment $\mathbf{D}$ in model $\mathbb{F}_1$ and let $\mathbf{U}^{(\mathbb{F}_2)}(t) = \{\mathbf{S}, \mathbf{I}, \mathbf{D}, \mathbf{R}, \mathbf{I}^c\}$. The governing eqaution of the dynamics takes the form $\frac{(\Delta t)^{\vec{\kappa}-1}}{\Gamma(\vec{\kappa}+1)}\, {}_0^C\mathscr{D}_t^{\vec{\kappa}(t)} \mathbf{U}^{(\mathbb{F}_2)}(t) = \mathscr{F}^{(\mathbb{F}_2)}(\mathbf{U}^{(\mathbb{F}_2)}, t; \lambda)$; see Table 3 second column.

**Table 2.** Integer-order models. Definitions and values/ranges for parameters are given in Table 1.

| Model ($\mathbb{I}_1$) | Model ($\mathbb{I}_2$) | Model ($\mathbb{I}_3$) |
|---|---|---|
| $\frac{d}{dt}\mathbf{S} = -\frac{\beta_I[\mathbf{I}+\varepsilon\mathbf{J}]}{N}\mathbf{S} - \frac{\nu}{N}\mathbf{S},$<br>$\frac{d}{dt}\mathbf{E} = \frac{\beta_I[\mathbf{I}+\varepsilon\mathbf{J}]}{N}\mathbf{S} - \alpha\mathbf{E},$<br>$\frac{d}{dt}\mathbf{I} = \delta\alpha\mathbf{E} - \gamma\mathbf{I},$<br>$\frac{d}{dt}\mathbf{J} = (1-\delta)\alpha\mathbf{E} - \gamma_a\mathbf{J},$<br>$\frac{d}{dt}\mathbf{D} = q\phi_D\mathbf{H},$<br>$\frac{d}{dt}\mathbf{H} = p\gamma\mathbf{I} - q\phi_D\mathbf{H} -(1-q)\phi_R\mathbf{H},$<br>$\frac{d}{dt}\mathbf{R} = \gamma_a\mathbf{J} + (1-p)\gamma\mathbf{I} +(1-q)\phi_R\mathbf{H} + \frac{\nu}{N}\mathbf{S},$<br>$\frac{d}{dt}\mathbf{I}^c = \delta\alpha\mathbf{E},$<br>$\frac{d}{dt}\mathbf{H}^c = p\gamma\mathbf{I},$ | $\frac{d}{dt}\mathbf{S} = -\frac{\beta_I[\chi\mathbf{P}+\mathbf{I}+\varepsilon\mathbf{J}]}{N}\mathbf{S} - \frac{\nu}{N}\mathbf{S},$<br>$\frac{d}{dt}\mathbf{E} = \frac{\beta_I[\chi\mathbf{P}+\mathbf{I}+\varepsilon\mathbf{J}]}{N}S - \alpha_1\mathbf{E},$<br>$\frac{d}{dt}\mathbf{P} = \alpha_1\mathbf{E} - \alpha_2\mathbf{P},$<br>$\frac{d}{dt}\mathbf{I} = \delta\alpha_2\mathbf{P} - \gamma\mathbf{I},$<br>$\frac{d}{dt}\mathbf{J} = (1-\delta)\alpha_2\mathbf{P} - \gamma_a\mathbf{J},$<br>$\frac{d}{dt}\mathbf{D} = q\phi_D\mathbf{H},$<br>$\frac{d}{dt}\mathbf{H} = p\gamma\mathbf{I} - q\phi_D\mathbf{H} -(1-q)\phi_R\mathbf{H},$<br>$\frac{d}{dt}\mathbf{R} = \gamma_a\mathbf{J} + (1-p)\gamma\mathbf{I} +(1-q)\phi_R\mathbf{H} + \frac{\nu}{N}\mathbf{S},$<br>$\frac{d}{dt}\mathbf{I}^c = \delta\alpha_2\mathbf{P},$<br>$\frac{d}{dt}\mathbf{H}^c = p\gamma\mathbf{I},$ | $\frac{d}{dt}\mathbf{S} = -\frac{\beta_I[\mathbf{I}+\varepsilon\mathbf{P}+\varepsilon\mathbf{J}]}{N}\mathbf{S} - \frac{\nu}{N}\mathbf{S},$<br>$\frac{d}{dt}\mathbf{E} = \frac{\beta_I[\mathbf{I}+\varepsilon\mathbf{P}+\varepsilon\mathbf{J}]}{N}\mathbf{S} - \alpha_1\mathbf{E},$<br>$\frac{d}{dt}\mathbf{P} = \alpha_1\mathbf{E} - \alpha_2\mathbf{P},$<br>$\frac{d}{dt}\mathbf{I} = \delta\alpha_2\mathbf{P} - d_I\mathbf{I},$<br>$\frac{d}{dt}\mathbf{J} = (1-\delta)\alpha_2\mathbf{P} - \gamma_a\mathbf{J},$<br>$\frac{d}{dt}\mathbf{D} = qd_H\mathbf{H},$<br>$\frac{d}{dt}\mathbf{H} = pd_I\mathbf{I} - d_H\mathbf{H},$<br>$\frac{d}{dt}\mathbf{Q} = (1-p)d_I\mathbf{I} - d_Q\mathbf{Q},$<br>$\frac{d}{dt}\mathbf{R} = \gamma_a\mathbf{J} + (1-q)d_H\mathbf{H} +d_Q\mathbf{Q} + \frac{\nu}{N}\mathbf{S},$<br>$\frac{d}{dt}\mathbf{I}^c = \delta\alpha_2\mathbf{P},$<br>$\frac{d}{dt}\mathbf{H}^c = pd_I\mathbf{I},$ |

**Table 3.** Fractional-order models. Definitions and values/ranges for parameters are given in Table 1.

| Model ($\mathbb{F}_1$) | Model ($\mathbb{F}_2$) | Model ($\mathbb{F}_3$) |
|---|---|---|
| $\frac{(\Delta t)^{\kappa_1-1}}{\Gamma(\kappa_1+1)}\,{}_0^C\mathscr{D}_t^{\kappa_1}\mathbf{S} = -\frac{\beta_I}{N}\mathbf{IS},$<br>$\frac{(\Delta t)^{\kappa_2-1}}{\Gamma(\kappa_2+1)}\,{}_0^C\mathscr{D}_t^{\kappa_2}\mathbf{I} = \frac{\beta_I}{N}\mathbf{IS} - \gamma_1\mathbf{I},$<br>$\frac{(\Delta t)^{\kappa_3-1}}{\Gamma(\kappa_3+1)}\,{}_0^C\mathscr{D}_t^{\kappa_3}\mathbf{R} = \gamma_1\mathbf{I},$<br>$\frac{(\Delta t)^{\kappa_2-1}}{\Gamma(\kappa_2+1)}\,{}_0^C\mathscr{D}_t^{\kappa_2}\mathbf{I}^c = \frac{\beta_I}{N}\mathbf{IS}.$ | $\frac{(\Delta t)^{\kappa_1(t)-1}}{\Gamma(\kappa_1(t)+1)}\,{}_0^C\mathscr{D}_t^{\kappa_1(t)}\mathbf{S} = -r\mathbf{IS},$<br>$\frac{(\Delta t)^{\kappa_2(t)-1}}{\Gamma(\kappa_2(t)+1)}\,{}_0^C\mathscr{D}_t^{\kappa_2(t)}\mathbf{I} = r\mathbf{IS} - (a+b)\mathbf{I},$<br>$\frac{(\Delta t)^{\kappa_3(t)-1}}{\Gamma(\kappa_3(t)+1)}\,{}_0^C\mathscr{D}_t^{\kappa_3(t)}\mathbf{D} = b\mathbf{I},$<br>$\frac{(\Delta t)^{\kappa_4(t)-1}}{\Gamma(\kappa_4(t)+1)}\,{}_0^C\mathscr{D}_t^{\kappa_4(t)}\mathbf{R} = a\mathbf{I}$<br>$\frac{(\Delta t)^{\kappa_2(t)-1}}{\Gamma(\kappa_2(t)+1)}\,{}_0^C\mathscr{D}_t^{\kappa_2(t)}\mathbf{I}^c = r\mathbf{IS}.$ | $\frac{(\Delta t)^{\kappa_1(t)-1}}{\Gamma(\kappa_1(t)+1)}\,{}_0^C\mathscr{D}_t^{\kappa_1(t)}\mathbf{S} = -\frac{\beta_I}{N}\mathbf{IS},$<br>$\frac{(\Delta t)^{\kappa_2(t)-1}}{\Gamma(\kappa_2(t)+1)}\,{}_0^C\mathscr{D}_t^{\kappa_2(t)}\mathbf{I} = \frac{\beta_I}{N}\mathbf{IS} - \gamma\mathbf{I},$<br>$\frac{(\Delta t)^{\kappa_3(t)-1}}{\Gamma(\kappa_3(t)+1)}\,{}_0^C\mathscr{D}_t^{\kappa_3(t)}\mathbf{H} = p\gamma\mathbf{I} - q\phi_D\mathbf{H} -(1-q)\phi_R\mathbf{H},$<br>$\frac{(\Delta t)^{\kappa_4(t)-1}}{\Gamma(\kappa_4(t)+1)}\,{}_0^C\mathscr{D}_t^{\kappa_4(t)}\mathbf{D} = q\phi_D\mathbf{H},$<br>$\frac{(\Delta t)^{\kappa_5(t)-1}}{\Gamma(\kappa_5(t)+1)}\,{}_0^C\mathscr{D}_t^{\kappa_5(t)}\mathbf{R} = (1-p)\gamma\mathbf{I} +(1-q)\phi_R\mathbf{H},$<br>$\frac{(\Delta t)^{\kappa_2(t)-1}}{\Gamma(\kappa_2(t)+1)}\,{}_0^C\mathscr{D}_t^{\kappa_2(t)}\mathbf{I}^c = \frac{\beta_I}{N}\mathbf{IS},$<br>$\frac{(\Delta t)^{\kappa_3(t)-1}}{\Gamma(\kappa_3(t)+1)}\,{}_0^C\mathscr{D}_t^{\kappa_3(t)}\mathbf{H}^c = p\gamma\mathbf{I}.$ |

**Table 4.** Time-delay model. Definitions and values/ranges for parameters are given in Table 1. $\lambda(t) = \frac{\beta_I[\mathbf{I}+\varepsilon\mathbf{J}]}{N}$.

| Model ($\mathbb{T}_1$) |
| --- |
| $\frac{\mathrm{d}}{\mathrm{d}t}\mathbf{S} = -\lambda(t-d)\mathbf{S}(t-d),$ |
| $\frac{\mathrm{d}}{\mathrm{d}t}\mathbf{I} = \delta\lambda(t-d)\mathbf{S}(t-d) - \gamma\mathbf{I},$ |
| $\frac{\mathrm{d}}{\mathrm{d}t}\mathbf{J} = (1-\delta)\lambda(t-d)\mathbf{S}(t-d) - \gamma_a\mathbf{J},$ |
| $\frac{\mathrm{d}}{\mathrm{d}t}\mathbf{H} = p\gamma\mathbf{I} - d_H\mathbf{H},$ |
| $\frac{\mathrm{d}}{\mathrm{d}t}\mathbf{D} = qd_H\mathbf{H},$ |
| $\frac{\mathrm{d}}{\mathrm{d}t}\mathbf{R} = \gamma_a\mathbf{J} + (1-p)\gamma\mathbf{I} + (1-q)d_H\mathbf{H},$ |
| $\frac{\mathrm{d}}{\mathrm{d}t}\mathbf{I}^c = \delta\lambda(t-d)\mathbf{S}(t-d),$ |
| $\frac{\mathrm{d}}{\mathrm{d}t}\mathbf{H}^c = p\gamma\mathbf{I},$ |

Here, $\vec{\kappa}(t) = (\kappa_1(t), \kappa_2(t), \kappa_3(t), \kappa_4(t)) \in (0,1)^4$ with $\kappa_1(t)$, $\kappa_2(t)$, $\kappa_3(t)$, and $\kappa_4(t)$ being the fractional derivative orders for the compartments **S**, **I**, **D**, and **R**, respectively. The effective reproduction number is given by $\mathscr{R}_c = \frac{r}{b}$.

• Model $\mathbb{F}_3$: Fractional-order SIHDR. We include two additional compartments **H**, **D** into the fractional-order model ($\mathbb{F}_1$). Therefore, we let $\mathbf{U}^{(\mathbb{F}_3)} = \{\mathbf{S}(t), \mathbf{I}(t), \mathbf{H}(t), \mathbf{D}(t), \mathbf{R}(t), \mathbf{I}^c(t), \mathbf{H}^c(t)\}$. The governing equation of the dynamics takes the form $\frac{(\Delta t)^{\vec{\kappa}(t)-1}}{\Gamma(\vec{\kappa}(t)+1)} {}_0^C\mathscr{D}_t^{\vec{\kappa}(t)}\mathbf{U}^{(\mathbb{F}_3)}(t) = \mathscr{F}^{(\mathbb{F}_3)}(\mathbf{U}^{(\mathbb{F}_3)}, t; \lambda)$; see Table 3 third column. Here, $\vec{\kappa}(t) = (\kappa_1(t), \kappa_2(t), \kappa_3(t), \kappa_4(t), \kappa_5(t)) \in (0,1)^5$ with $\kappa_1(t)$, $\kappa_2(t)$, $\kappa_3(t)$, $\kappa_4(t)$, and $\kappa_5(t)$ being the fractional derivative orders for the compartments **S**, **I**, **H**, **D**, and **R**, respectively. The effective reproduction number is given by $\mathscr{R}_c = \frac{\beta_I}{\gamma}$.

#### 3.4 The Corresponding Integer-Order Models of Fractional-Order Models $\mathbb{F}_1$ and $\mathbb{F}_3$

In the identifiability analysis of fractional-order models $\mathbb{F}_1$ and $\mathbb{F}_3$ (discussed in Section 4 in Supplementary and Results section in the main text), we use the simulation of the corresponding integer-order models. Here, we provide definitions of these models and name them models $\mathbb{I}_4$ and $\mathbb{I}_5$, respectively.

• Model $\mathbb{I}_4$: Integer-order SIR. We consider the corresponding integer-order model of model $\mathbb{F}_1$. We let $\mathbf{U}^{(\mathbb{I}_4)}(t) = \{\mathbf{S}, \mathbf{I}, \mathbf{R}, \mathbf{I}^c\}$ and thus the governing equation of the dynamics takes the form $\frac{\mathrm{d}}{\mathrm{d}t}\mathbf{U}^{(\mathbb{I}_4)}(t) = \mathscr{F}^{(\mathbb{I}_4)}(\mathbf{U}^{(\mathbb{I}_4)}, t; \lambda)$, which is the same equations as in Table 3 first column, where $\kappa_1 = \kappa_2 = \kappa_3 = 1$.

• Model $\mathbb{I}_5$: Integer-order SIHDR. We consider the corresponding integer-order model of model $\mathbb{F}_3$. We let $\mathbf{U}^{(\mathbb{I}_5)}(t) = \{\mathbf{S}, \mathbf{I}, \mathbf{H}, \mathbf{D}, \mathbf{R}, \mathbf{I}^c\}$. The governing equation of the dynamics takes the form $\frac{\mathrm{d}}{\mathrm{d}t}\mathbf{U}^{(\mathbb{I}_5)}(t) = \mathscr{F}^{(\mathbb{I}_5)}(\mathbf{U}^{(\mathbb{I}_5)}, t; \lambda)$, which is the same equations as in Table 3 third column, where $\kappa_1(t) = \kappa_2(t) = \kappa_3(t) = \kappa_4(t) = \kappa_5(t) = 1$.

### 4 Detailed discussion on different sets of results

In this section, we show the results of integer-order models $\mathbb{I}_1$, $\mathbb{I}_2$, $\mathbb{I}_3$, and time-delay model $\mathbb{T}_1$ based on MI data[17,18] and RI data[19]. We also show the simulation results of fractional-order model $\mathbb{F}_1$ based on NYC data[20].

#### 4.1 Michigan (MI) Data Set

We simulate models $\mathbb{I}_1$, $\mathbb{I}_2$, $\mathbb{I}_3$, and the time-delay model $\mathbb{T}_1$ based on the MI data set[17]. The training data set consists of the daily infectious cases $\mathbf{I}^{new}$, current hospitalized cases **H**, and the daily death cases $\mathbf{D}^{new}$ since April 15, 2020. In this case, the parameter $q(t)$ can be calculated by $q(t) = \phi_D \frac{\mathbf{D}^{new}(t)}{\mathbf{H}(t)}$, $t \in \mathbb{N}$. In the PINN formulation, we use a neural network of 10 hidden layers with 32 neurons per layer and tanh activation function to approximate $\mathbf{U}(t)$. The time-dependent parameters are parameterized by separate networks of 5 hidden layers with 20 neurons per layer and tanh activation function. Figure 2 shows the simulation results based on MI data set. Panel **A** shows the accurate fitting to the available data. Panel **B** and Panel **C** compare the inference of the time-dependent parameters and unobserved dynamics, respectively. Based on the inferred parameters, we show the prediction of the dynamics in Panel **D**. Here, we approximate the effective vaccination per day ($V(t)$) by the following piece-wise linear function

$$V(t) = \begin{cases} 0, & 0 \le t < 260, \\ 500(t-300), & 260 \le t < 300, \\ 20000, & t \ge 300, \end{cases} \tag{6}$$

after post-processing the vaccination data of MI[18].

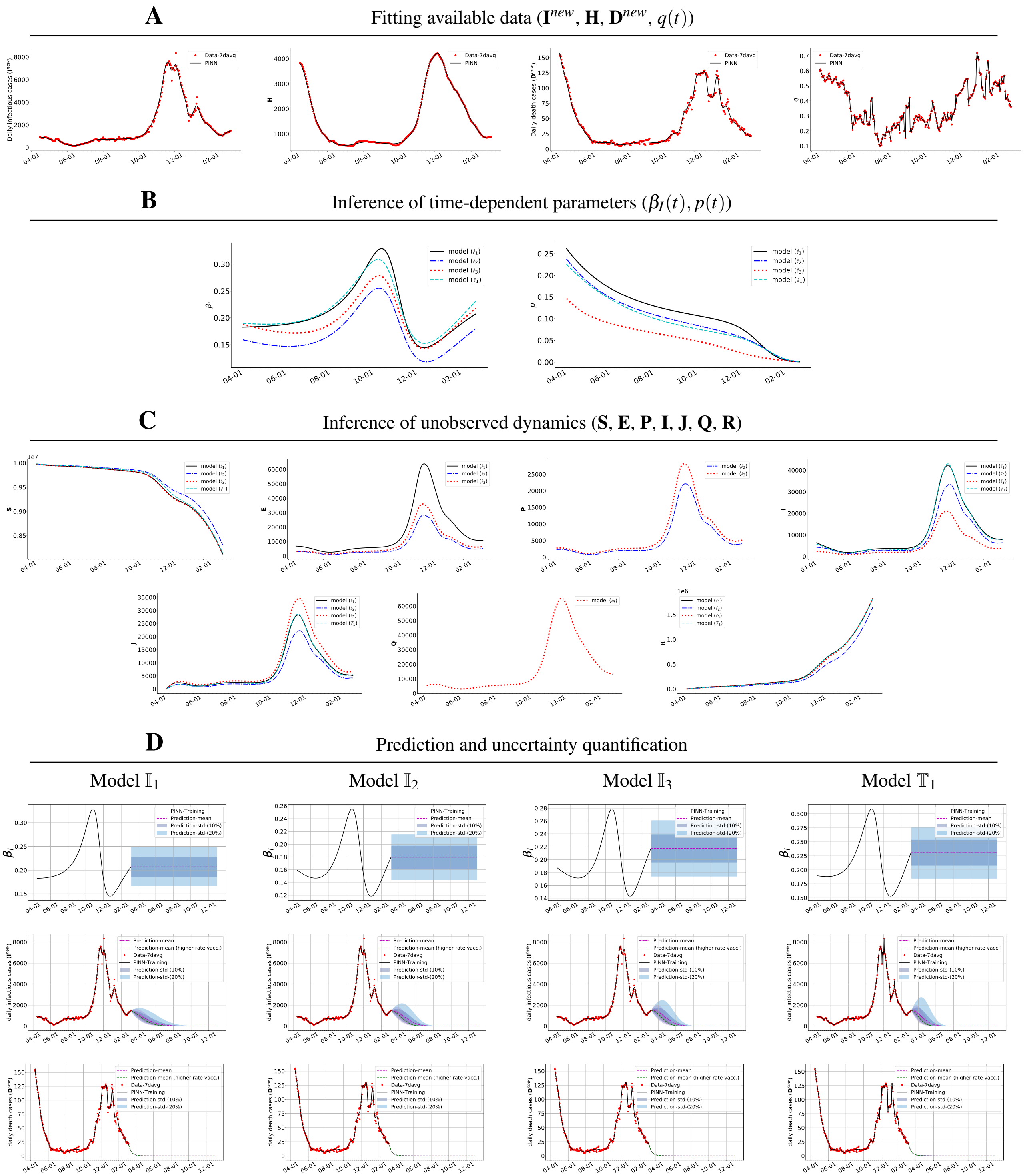


**Figure 2. PINNs inference using the integer-order models $\mathbb{I}_1$, $\mathbb{I}_2$, $\mathbb{I}_3$ and time-delay model $\mathbb{T}_1$ for MI**. **A**: Accurate fitting to the available data of daily infectious and death cases and the current hospitalized cases. **B**: Inference of time-dependent parameters ($\beta_I(t)$, $p(t)$). **C**: Inference of unobserved dynamics. **D**: Prediction and uncertainty quantification of daily infectious and death cases. Here the inferred delay is $d = 3.21$ for the time-delay model $\mathbb{T}_1$.

#### 4.2 Rhode Island (RI) Data Set

We simulate models $\mathbb{I}_1$, $\mathbb{I}_2$, $\mathbb{I}_3$, and the time-delay model $\mathbb{T}_1$ based on RI data set[19]. The training data set includes daily infectious cases $\mathbf{I}^{new}$, daily hospitalized cases $\mathbf{H}^{new}$, current hospitalized cases $\mathbf{H}$, and the daily death cases $\mathbf{D}^{new}$ since March 04, 2020. In this case, the parameter $q(t)$ can be calculated by $q(t) = \phi_D \frac{\mathbf{D}^{new}(t)}{\mathbf{H}(t)}$, $t \in \mathbb{N}$. The structure of neural network is the same as in the MI dataset. Figure 3 shows the simulation results based on RI data set. Panel **A** shows the accurate fitting to the available data. Panel **B** and Panel **C** compare the inference of the time-dependent parameters and unobserved dynamics, respectively. Based on the inferred parameters, we show the prediction of the dynamics in Panel **D**. Here, we approximate the effective vaccination per day ($V(t)$) by the following piece-wise linear function

$$V(t) = \begin{cases} 0, & 0 \le t < 300, \\ 47.72(t-300)+368, & 300 \le t < 356, \\ 95.43(Dt-356)+3408, & 356 \le t < 600, \\ 7607, & t \ge 600, \end{cases} \tag{7}$$

after post-processing the vaccination data of RI.

#### 4.3 NYC Data Set

We discuss the identifiability of fractional model $\mathbb{F}_1$ based on the NYC data[20] and show the results in Fig. 4. As a comparison, we plot the results of fractional model $\mathbb{F}_1$ against the results of its corresponding integer-order model $\mathbb{I}_4$ with time-dependent parameter. In the first three rows of each column, black color shows the results of model $\mathbb{F}_1$ and red color shows the results of corresponding integer-order model $\mathbb{I}_4$. The first column shows the inferred dynamics and fractional orders $\kappa_i$, $i = 1, 2, 3$, based on the only available data $\mathbf{I}^{new}$. Although with a small uncertainty bound, the inferred compartment $\mathbf{I}$ shows an erroneous trend with a sharp increase at the end of the training. The uncertainty bounds for $\mathbf{S}$, $\mathbf{R}$, and the fractional orders in this case are also larger compared to the other cases due to lack of data. In the second column, we assume that in addition to daily infectious cases $\mathbf{I}^{new}$, the compartment $\mathbf{S}$ is available from the corresponding integer-order model. It can be observed that the integer-order model results cannot be recovered even though the fractional orders converge to 1 in this case. In the third column, we assume that in addition to the daily infectious cases $\mathbf{I}^{new}$, the compartment $\mathbf{I}$ is available from the corresponding integer-order model. In the last column, we assume that in addition to the $\mathbf{I}^{new}$, the compartments $\mathbf{S}$, $\mathbf{I}$, and $\mathbf{R}$ are available. We observer that the uncertainty bounds are reduced when we have more data.

### 5 Detailed formulation of PINNs for integer- and fractional-order models

In this section, we explain the PINN formulations that we use to infer the unobserved dynamics and time-dependent parameters of the nine different models that we consider.

#### 5.1 Physics-Informed Neural Networks (PINNs)

The PINN formulation has been recently developed in[21], which constructs a physics-informed deep learning algorithm to solve the forward and inverse problems involving differential equations by employing a deep neural network to approximate the unknown function. In particular in our study, we let $\mathbf{U}_{NN}(t;\Theta)$ be a deep neural network with input $t$, which is parameterized by $\Theta$ as weights and biases of the network; see Fig. 5 the green-shaded part. We approximate the solution of differential equation by the neural network, i.e. $\mathbf{U}(t) \approx \mathbf{U}_{NN}(t;\Theta)$. We define the residual of equation as $\mathscr{R}_{NN}(t) = \frac{\mathrm{d}}{\mathrm{d}t}\mathbf{U}_{NN}(t) - \mathscr{F}(\mathbf{U}_{NN}, t; \lambda)$ and encode this residual into the network; see the orange-shaded area in Fig. 5. The unknown time-dependent parameters in the governing equation are also parametrized by separate neural networks; see the red-shaded area in Fig. 5. In the PINN formulation, we define two finite sets of training points $\{t_u^j\}_{j=1}^{N_u}$ and residual points $\{t_r^j\}_{j=1}^{N_r}$. The training points are the points where we have the data available and the residual points are the points where the residual $\mathscr{R}_{NN}(t)$ is satisfied and they are randomly selected over the entire computational domain. Therefore, we define the loss function of PINN as

$$L(\Theta,\lambda) = \omega_u \mathrm{MSE}_u + \omega_r \mathrm{MSE}_r = \omega_u \frac{1}{N_u} \sum_{j=1}^{N_u} \left| \mathbf{U}_{NN}(t_u^j) - \mathbf{U}^{(\mathbb{D})}(t_u^j) \right|^2 + \omega_r \frac{1}{N_r} \sum_{j=1}^{N_r} \left| \mathscr{R}_{NN}(t_r^j) \right|^2, \tag{8}$$

where MSE stands for mean squared error. We see that the loss function of PINN contains two terms. The $\mathrm{MSE}_u$ measures the mismatch between solution $\mathbf{U}_{NN}$ and data $\mathbf{U}^{(\mathbb{D})}$ at the training points, which depends on the availability of data on the epidemiological classes as discussed in Section 5.3. The $\mathrm{MSE}_r$ penalizes the governing equation at the residual points. We provide the detailed expansion of this term in Section 5.4 equation (15) for model $\mathbb{I}_1$. Similar expansions can be obtained for other integer-order models $\mathbb{I}_1$, $\mathbb{I}_2$, $\mathbb{I}_3$, and time-delay model $\mathbb{T}_1$. In each model, we denote the output of the network as $\mathbf{U}_{NN}^{(\mathbb{I}_1)}(t)$, $\mathbf{U}_{NN}^{(\mathbb{I}_2)}(t)$, $\mathbf{U}_{NN}^{(\mathbb{I}_3)}(t)$, and $\mathbf{U}_{NN}^{(\mathbb{T}_1)}(t)$, respectively. We consider three separate networks to represent the time-dependent parameters

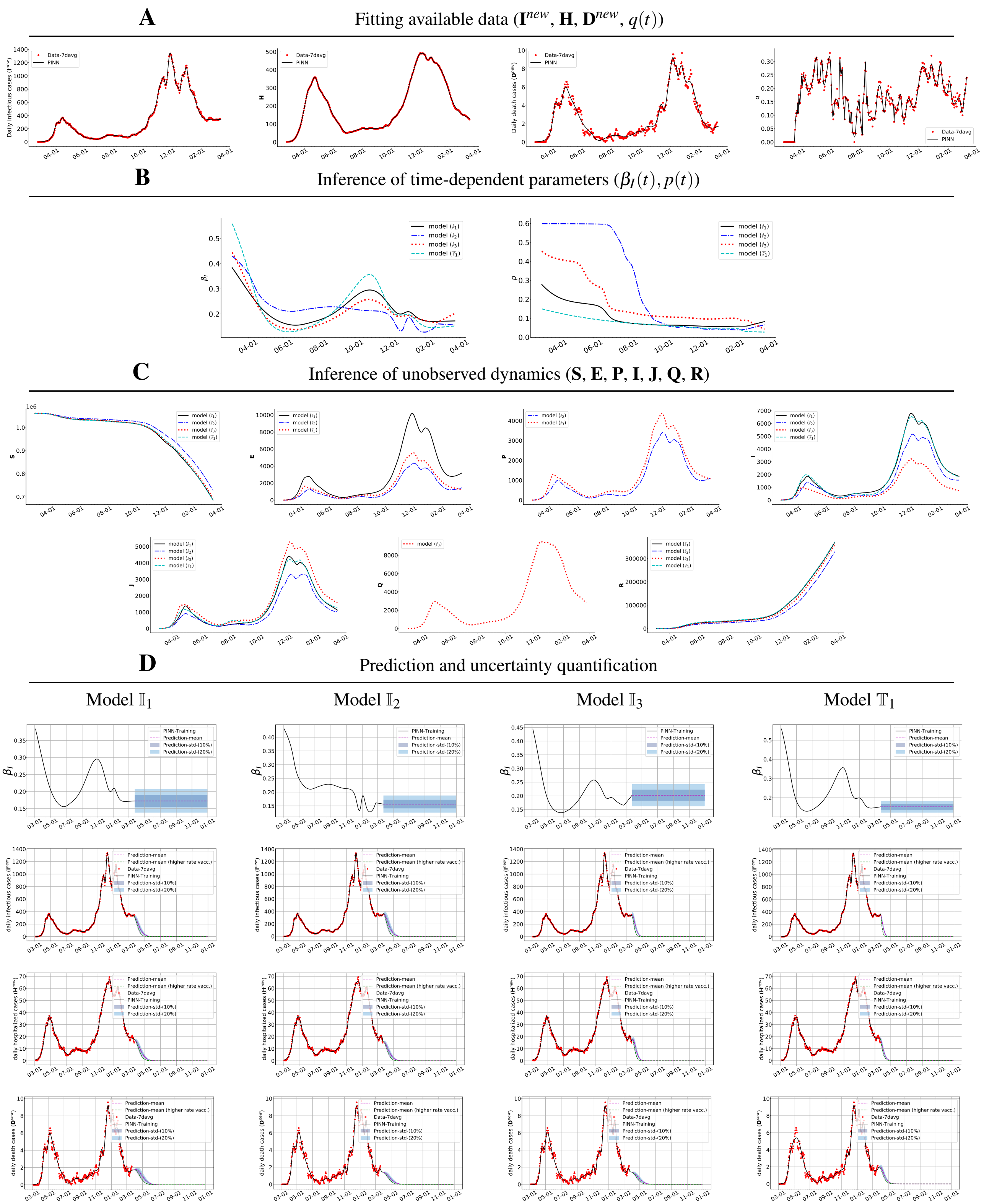


**Figure 3.** **PINNs inference using the integer-order models $\mathbb{I}_1$, $\mathbb{I}_2$, $\mathbb{I}_3$ and time-delay model $\mathbb{T}_1$ for RI**. **A**: Accurate fitting to the available data of daily infectious and death cases and the current hospitalized cases. **B**: Inference of time-dependent parameters ($\beta_I(t)$, $p(t)$). **C**: Inference of unobserved dynamics. **D**: Prediction and uncertainty quantification of daily infectious, hospitalized, and death cases. Here the inferred delay is $d = 8.97$ for the time-delay model $\mathbb{T}_1$.

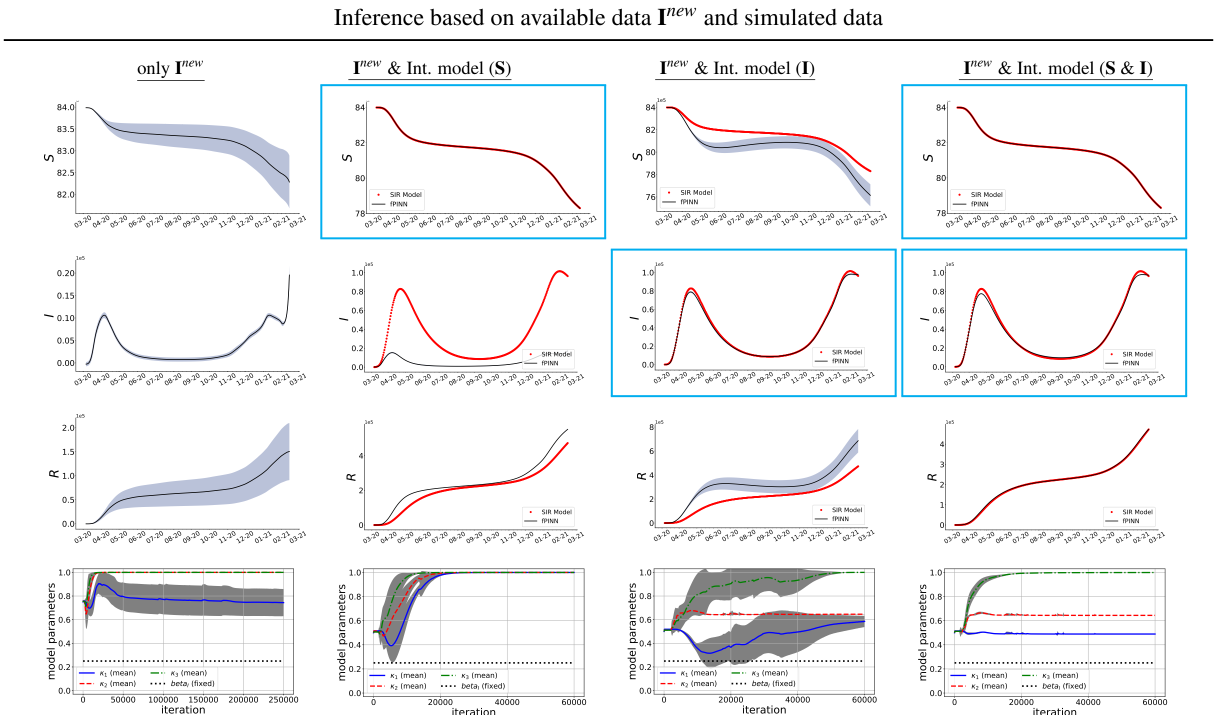


**Figure 4. Identifiability study for PINNs for the fractional order model $\mathbb{F}_1$ for NYC**. The parameters $\beta = 0.25$ and $\gamma = 0.0365$ are fixed in the fractional model. Instead of time-dependent parameters, here we aim to infer unobserved dynamics and different fractional operators for $\kappa_i$, $i = 1, 2, 3$. First column: Inference based on only available data $\mathbf{I}^{new}$; Second column: Inference based on available data $\mathbf{I}^{new}$ and $\mathbf{S}$ from the corresponding integer-order SIR model; Third column: Inference based on available data $\mathbf{I}^{new}$ and $\mathbf{I}$ from the corresponding integer-order SIR model; Fourth column: Inference based on available data $\mathbf{I}^{new}$, $\mathbf{S}$, $\mathbf{I}$, and $\mathbf{R}$ from the corresponding integer-order SIR model.

$\beta_I(t)$, $p(t)$ and $q(t)$. In time-delay model, we use a separate network to represent the time-delay parameter. In all cases, the derivatives of network with respect to the input $t$ and all network parameters $\Theta$ are computed by applying the chain rule for differentiating compositions of functions using the automatic differentiation[22]. In particular, we use Tensorflow programming[23], which is a popular and relatively well documented open source software library for automatic differentiation and deep learning computations.

### 5.2 Fractional Physics-Informed Neural Networks (fPINNs)

Fractional PINNs (fPINNs)[24] extend PINNs to solve forward and inverse problems with fractional differential operators. Since the standard chain rule for integer-order derivatives is not valid for the fractional ones, fPINN formulation does not use automatic differentiation to compute the fractional derivatives. Instead, the residual in the loss function of fPINNs adopts a numerical discretization for the fractional operators. This will require additional set of "auxiliary" points to numerically compute the fractional derivatives. In particular, the Caputo fractional derivative used in this paper is numerically approximated by the L1 scheme[25,26] on a uniform mesh $\{t_n = n\tau\}$ as ,

$$ {}_0^C\mathscr{D}_t^{\kappa(t)} f(t_n) = \delta_t^{\kappa(t)} f(t_n) \triangleq \sum_{k=0}^{n-1} b_{n,n-k-1}[f(t_{k+1}) - f(t_k)] + \mathscr{O}(\tau^{2-\kappa(t)}), \tag{9} $$

where $\kappa(t) \in (0,1)$, $\tau$ is the (sufficiently small) time step, and the coefficients are given by $b_{n,k} = \frac{\tau^{-\kappa(t_n)}}{\Gamma(2-\kappa(t_t))}[(k+1)^{1-\kappa(t_n)} - k^{1-\kappa(t_n)}]$. The formulation of fPINNs is very similar to PINNs. We let $\mathbf{U}_{NN}^{(\mathbb{F})}(t;\Theta)$ be a deep neural network with input $t$, parameterized by $\Theta$ as weights and biases of the network. We approximate the solution of fractional model by $\mathbf{U}(t) \approx \mathbf{U}_{NN}^{(\mathbb{F})}(t;\Theta)$ and define the residual as

$$ \mathscr{R}_{NN}^{(\mathbb{F})}(t) = \frac{(\Delta t)^{\kappa(t)-1}}{\Gamma(\kappa(t)+1)} {}_0^C\mathscr{D}_t^{\kappa(t)} \mathbf{U}_{NN}^{(\mathbb{F})}(t) - \mathscr{F}^{(\mathbb{F})}(\mathbf{U}_{NN}^{(\mathbb{F})}, t; \lambda) \approx \frac{(\Delta t)^{\kappa(t)-1}}{\Gamma(\kappa(t)+1)} \delta_t^{\kappa(t)} \mathbf{U}_{NN}^{(\mathbb{F})}(t) - \mathscr{F}^{(\mathbb{F})}(\mathbf{U}_{NN}^{(\mathbb{F})}, t; \lambda). \tag{10} $$

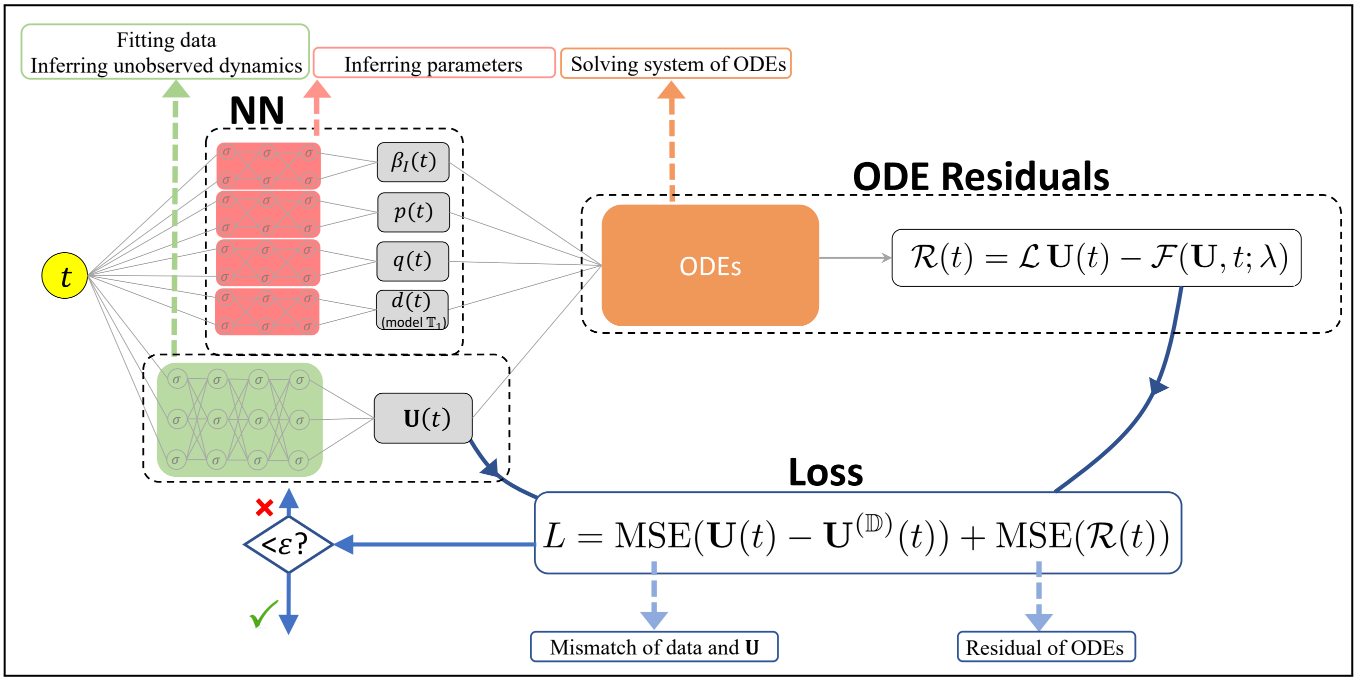


**Figure 5. Schematic of physics-informed neural networks**. **NN** denotes different neural networks representing the states $\mathbf{U}(t)$ (green-shaded area) and the model parameters $\beta_I(t)$, $p(t)$, $q(t)$ in integer-order models and $d$ in time-delay model (red-shaded area). **ODE Residuals**: computes the residual of models. **Loss**: is comprised of both terms from the mismatch between data and NN and the ODE residuals.

Similar to PINNs, in the fPINN formulation, we define two finite sets of training points $\{t_u^j\}_{j=1}^{N_u}$ and residual points $\{t_r^j\}_{j=1}^{N_r}$. The training points are the points where we have the data available and the residual points are the points where the residual (10) is satisfied and they are freely available all over the computational domain. We define the loss function of fPINN as

$$L^{(\mathbb{F})}(\Theta, \lambda) = \omega_u \text{MSE}_u^{(\mathbb{F})} + \omega_r \text{MSE}_r^{(\mathbb{F})} = \omega_u \frac{1}{N_u} \sum_{j=1}^{N_u} \left| \mathbf{U}_{NN}^{(\mathbb{F})}(t_u^j) - \mathbf{U}^{(\mathbb{D})}(t_u^j) \right|^2 + \omega_r \frac{1}{N_r} \sum_{j=1}^{N_r} \left| \mathscr{R}_{NN}^{(\mathbb{F})}(t_r^j) \right|^2. \tag{11}$$

where MSE stands for mean squared error. The $\text{MSE}_u$ is defined similar to PINNs. The detailed expansions for $\text{MSE}_r^{(\mathbb{F})}$ for different fractional models are given in Section 5.4. We note that in fPINNs, the number of residual points depends on the the time step $\tau$. While a small time step improves the discretization error, it increases the number of residual points, which will further impose extra computational costs. The computational bottleneck is the for loop in computing the fractional derivative. We show in Sec. 5.5 a vectorization technique that we implemented to reduce the computational cost.

We apply fPINNs to solve the fractional models $\mathbb{F}_1$, $\mathbb{F}_2$, and $\mathbb{F}_3$ with the network output denoted by $\mathbf{U}_{NN}^{(\mathbb{F}_1)}(t)$, $\mathbf{U}_{NN}^{(\mathbb{F}_2)}(t)$, and $\mathbf{U}_{NN}^{(\mathbb{F}_3)}(t)$, respectively. In the fractional models, we fix the model parameters and infer the fractional order $\kappa_i(t)$'s, as well as the unobserved dynamics. We use a separate network to represent each time-dependent fractional order.

### 5.3 Training Data

Not all of the compartments in an epidemiological model are tractable in practice. Hence, the reported data of individuals is restricted to only a few compartments. Here, in this study, we have access to two categories of reported datasets. They both include the history of the cumulative death $\mathbf{D}^c(t)$ (note that since death compartment does not have an outflow then the current and cumulative values are the same, i.e., $\mathbf{D}^c(t) = \mathbf{D}(t)$). Set 1, includes cumulative infectious $\mathbf{I}^c(t)$, cumulative hospitalized individuals $\mathbf{H}^c(t)$, and may also include the current hospitalized individuals $\mathbf{H}(t)$. Set 2 includes the current values of the infectious individuals $\mathbf{I}(t)$ and recovered individuals $\mathbf{R}(t)$. Thus,

- Set 1: $\mathbf{D}^c(t)$, $\mathbf{I}^c(t)$, $\mathbf{H}^c(t)$ (and/or $\mathbf{H}(t)$)
- Set 2: $\mathbf{D}^c(t)$, $\mathbf{I}(t)$, $\mathbf{R}(t)$

Given the cumulative values, we can obtain the daily increases for infectious, hospitalized, and death by $\mathbf{I}^{new}(t) = \mathbf{I}^c(t) - \mathbf{I}^c(t-1)$, $\mathbf{H}^{new}(t) = \mathbf{H}^c(t) - \mathbf{H}^c(t-1)$, and $\mathbf{D}^{new}(t) = \mathbf{D}^c(t) - \mathbf{D}^c(t-1)$. Based on the two categories of available data, the first term

$\text{MSE}_u$ in the loss functions (8) and (11) for the considered models has different formulations. For the data in Set 1, if only $\mathbf{H}^c(t)$ is given then the $\text{MSE}_u$ becomes

$$\begin{aligned}\text{MSE}_u = &\frac{1}{N_u}\sum_{j=1}^{N_u}|\mathbf{I}_{NN}^{new}(t_j) - I^{new}(t_j)|^2 + \frac{1}{N_u}\sum_{j=1}^{N_u}|\mathbf{H}_{NN}^{new}(t_j) - \mathbf{H}^{new}(t_j)|^2 + \frac{1}{N_u}\sum_{j=1}^{N_u}|\mathbf{D}_{NN}^{new}(t_j) - \mathbf{D}^{new}(t_j)|^2 \\ &+ \frac{1}{N_u}\sum_{j=1}^{N_u}|\mathbf{I}_{NN}^{c}(t_j) - \mathbf{I}^{c}(t_j)|^2 + \frac{1}{N_u}\sum_{j=1}^{N_u}|\mathbf{H}_{NN}^{c}(t_j) - \mathbf{H}^{c}(t_j)|^2 + \frac{1}{N_u}\sum_{j=1}^{N_u}|\mathbf{D}_{NN}^{c}(t_j) - \mathbf{D}^{c}(t_j)|^2.\end{aligned} \tag{12}$$

For the data in Set 1, if only $\mathbf{H}(t)$ is given then we have $q(t) = \phi_D \frac{\mathbf{D}^{new}(t)}{\mathbf{H}(t)}$ and then $\text{MSE}_u$ becomes

$$\begin{aligned}\text{MSE}_u = &\frac{1}{N_u}\sum_{j=1}^{N_u}|\mathbf{I}_{NN}^{new}(t_j) - I^{new}(t_j)|^2 + \frac{1}{N_u}\sum_{j=1}^{N_u}|\mathbf{D}_{NN}^{new}(t_j) - \mathbf{D}^{new}(t_j)|^2 \\ &+ \frac{1}{N_u}\sum_{j=1}^{N_u}|\mathbf{I}_{NN}^{c}(t_j) - \mathbf{I}^{c}(t_j)|^2 + \frac{1}{N_u}\sum_{j=1}^{N_u}|\mathbf{D}_{NN}^{c}(t_j) - \mathbf{D}^{c}(t_j)|^2 \\ &+ \frac{1}{N_u}\sum_{j=1}^{N_u}+|\mathbf{H}_{NN}(t_j) - \mathbf{H}(t_j)|^2 + \frac{1}{N_u}\sum_{j=1}^{N_u}|q_{NN}(t_j) - q(t_j)|^2\end{aligned} \tag{13}$$

For the data in Set 1, if both $\mathbf{H}^c(t)$ and $\mathbf{H}(t)$ are given then the $\text{MSE}_u$ becomes the combination of both (12) and (13). For the data in Set 2, the $\text{MSE}_u$ becomes

$$\text{MSE}_u = \frac{1}{N_u}\sum_{j=1}^{N_u}|\mathbf{I}_{NN}(t_j) - I(t_j)|^2 + \frac{1}{N_u}\sum_{j=1}^{N_u}|\mathbf{D}_{NN}^{c}(t_j) - \mathbf{D}^{c}(t_j)|^2 + \frac{1}{N_u}\sum_{j=1}^{N_u}|\mathbf{R}_{NN}(t_j) - \mathbf{R}(t_j)|^2. \tag{14}$$

In any case, if the data for one of the compartments is not available we omit the corresponding term from $\text{MSE}_u$.

#### 5.4 ODE Residuals

In view of the ODE system for model $\mathbb{I}_1$ in Table 2, the ODE residual for model $\mathbb{I}_1$ takes the form

$$\begin{aligned}\text{MSE}_r = &\frac{1}{N_f}\sum_{j=1}^{N_f}\left|\frac{\mathrm{d}}{\mathrm{d}t}\mathbf{E}_{NN}(t_j) - \frac{\beta_{INN}(t_j)\,[\mathbf{I}(t_j)+\varepsilon_1\mathbf{J}(t_j)]}{N}\mathbf{S}_{NN}(t_j) + \alpha\mathbf{E}_{NN}(t_j)\right|^2 \\ &+ \frac{1}{N_f}\sum_{j=1}^{N_f}\left|\frac{\mathrm{d}}{\mathrm{d}t}\mathbf{I}_{NN}(t_j) - \delta\alpha\mathbf{E}_{NN}(t_j) + \gamma\mathbf{I}_{NN}(t_j)\right|^2 + \frac{1}{N_f}\sum_{j=1}^{N_f}\left|\frac{\mathrm{d}}{\mathrm{d}t}\mathbf{D}_{NN}(t_j) - q_{NN}(t_j)\phi_D\mathbf{H}_{NN}(t_j)\right|^2 \\ &+ \frac{1}{N_f}\sum_{j=1}^{N_f}\left|\frac{\mathrm{d}}{\mathrm{d}t}\,\mathbf{J}_{NN}(t_j) - (1-\delta)\alpha\mathbf{E}_{NN}(t_j) + \gamma_a\mathbf{J}_{NN}(t_j)\right|^2 \\ &+ \frac{1}{N_f}\sum_{j=1}^{N_f}\left|\frac{\mathrm{d}}{\mathrm{d}t}\mathbf{H}_{NN}(t_j) - p_{NN}(t_j)\gamma\,\mathbf{I}_{NN}(t_j) + q_{NN}(t_j)\phi_D\mathbf{H}_{NN}(t_j) + (1-q_{NN}(t_j))\phi_R\mathbf{H}_{NN}(t_j)\right|^2 \\ &+ \frac{1}{N_f}\sum_{j=1}^{N_f}\left|\frac{\mathrm{d}}{\mathrm{d}t}\mathbf{R}_{NN}(t_j) - \gamma_a\,\mathbf{J}_{NN}(t_j) - (1-p_{NN}(t_j))\gamma\,\mathbf{I}_{NN}(t_j) - (1-q_{NN}(t_j))\phi_R\mathbf{H}_{NN}(t_j)\right|^2 \\ &+ \frac{1}{N_f}\sum_{j=1}^{N_f}\left|\frac{\mathrm{d}}{\mathrm{d}t}\mathbf{I}_{NN}^{c}(t_j) - \delta\alpha\mathbf{E}_{NN}(t_j)\right|^2 + \frac{1}{N_f}\sum_{j=1}^{N_f}\left|\frac{\mathrm{d}}{\mathrm{d}t}\,\mathbf{H}_{NN}^{c}(t_j) - p_{NN}(t_j)\gamma\,\mathbf{I}_{NN}(t_i)\right|^2\end{aligned} \tag{15}$$

with $\mathbf{S}_{NN}(t) = N - \mathbf{E}_{NN}(t) - \mathbf{I}_{NN}(t) - \mathbf{J}_{NN}(t) - \mathbf{D}_{NN}(t) - \mathbf{H}_{NN}(t) - \mathbf{R}_{NN}(t)$. Detailed expansions of the ODE residual for models $\mathbb{I}_2$, $\mathbb{I}_3$, and $\mathbb{T}_1$ can be similarly derived.

In view of the ODE systems for fractional models in Table 3, the ODE residual for model $\mathbb{F}_1$ takes the form

$$
\begin{aligned}
\mathrm{MSE}_r^{(\mathbb{F}_1)} = & \frac{1}{N_f}\sum_{j=1}^{N_f}\left|\frac{(\Delta t)^{\kappa_1-1}}{\Gamma(\kappa_1+1)}\delta_t^{\kappa_1}\mathbf{S}_{NN}(t_j)+\frac{\beta_I}{N}\mathbf{I}_{NN}(t_j)\mathbf{S}_{NN}(t_j)\right|^2 \\
& +\frac{1}{N_f}\sum_{j=1}^{N_f}\left|\frac{(\Delta t)^{\kappa_2-1}}{\Gamma(\kappa_2+1)}\delta_t^{\kappa_2}\mathbf{I}_{NN}(t_j)-\frac{\beta_I}{N}\mathbf{I}_{NN}(t_j)\mathbf{S}_{NN}(t_j)+\gamma_{1NN}(t_j)\mathbf{I}_{NN}(t_j)\right|^2 \\
& +\frac{1}{N_f}\sum_{j=1}^{N_f}\left|\frac{(\Delta t)^{\kappa_3-1}}{\Gamma(\kappa_3+1)}\delta_t^{\kappa_3}\mathbf{R}_{NN}(t_j)-\gamma_{1NN}(t_j)\;\mathbf{J}_{NN}\right|^2 \\
& +\frac{1}{N_f}\sum_{j=1}^{N_f}\left|\frac{(\Delta t)^{\kappa_2-1}}{\Gamma(\kappa_2+1)}\delta_t^{\kappa_2}\mathbf{I}^c_{NN}(t_j)-\frac{\beta_I}{N}\mathbf{I}_{NN}(t_j)\mathbf{S}_{NN}(t_j)\right|^2 \\
& +\frac{1}{N_f}\sum_{j=1}^{N_f}\left|N-\mathbf{S}_{NN}(t)-\mathbf{I}_{NN}(t)-\mathbf{R}_{NN}(t)\right|^2,
\end{aligned}
\tag{16}
$$

and the ODE residual for the fractional model $\mathbb{F}_3$ takes the form

$$
\begin{aligned}
\mathrm{MSE}_r^{(\mathbb{F}_3)} = & \frac{1}{N_f}\sum_{j=1}^{N_f}\left|\frac{(\Delta t)^{\kappa_{1_{NN}}(t_j)-1}}{\Gamma(\kappa_{1_{NN}}(t_j)+1)}\delta_t^{\kappa_{1_{NN}}(t_j)}\mathbf{S}_{NN}(t_j)+\frac{\beta_I}{N}\mathbf{I}_{NN}(t_j)\mathbf{S}_{NN}(t_j)\right|^2 \\
& +\frac{1}{N_f}\sum_{j=1}^{N_f}\left|\frac{(\Delta t)^{\kappa_{2_{NN}}(t_j)-1}}{\Gamma(\kappa_{2_{NN}}(t_j)+1)}\delta_t^{\kappa_{2_{NN}}(t_j)}\mathbf{I}_{NN}(t_j)-\frac{\beta_I}{N}\mathbf{I}_{NN}(t_j)\mathbf{S}_{NN}(t_j)+\gamma_{1NN}(t_j)\mathbf{I}_{NN}(t_j)\right|^2 \\
& +\frac{1}{N_f}\sum_{j=1}^{N_f}\left|\frac{(\Delta t)^{\kappa_{3_{NN}}(t_j)-1}}{\Gamma(\kappa_{3_{NN}}(t_j)+1)}\delta_t^{\kappa_{3_{NN}}(t_j)}\mathbf{H}_{NN}(t_j)-p_{NN}(t_j)\gamma\mathbf{I}_{NN}(t_j)+q_{NN}(t_j)\phi_D\mathbf{H}_{NN}(t_j)+(1-q_{NN}(t_j))\phi_R\mathbf{H}_{NN}(t_j)\right|^2 \\
& +\frac{1}{N_f}\sum_{j=1}^{N_f}\left|\frac{(\Delta t)^{\kappa_{4_{NN}}(t_j)-1}}{\Gamma(\kappa_{4_{NN}}(t_j)+1)}\delta_t^{\kappa_{4_{NN}}(t_j)}\mathbf{D}_{NN}(t_j)-q_{NN}(t_j)\phi_D\mathbf{H}_{NN}(t_j)\right|^2 \\
& +\frac{1}{N_f}\sum_{j=1}^{N_f}\left|\frac{(\Delta t)^{\kappa_{5_{NN}}(t_j)-1}}{\Gamma(\kappa_{5_{NN}}(t_j)+1)}\delta_t^{\kappa_{5_{NN}}(t_j)}\mathbf{R}_{NN}(t_j)-(1-p_{NN}(t_j))\gamma_{NN}(t_j)\mathbf{I}_{NN}-(1-q_{NN}(t_j))\phi_R\mathbf{H}_{NN}(t_j)\right|^2 \\
& +\frac{1}{N_f}\sum_{j=1}^{N_f}\left|\frac{(\Delta t)^{\kappa_{2_{NN}}(t_j)-1}}{\Gamma(\kappa_{2_{NN}}(t_j)+1)}\delta_t^{\kappa_{2_{NN}}(t_j)}\mathbf{I}^c_{NN}(t_j)-\frac{\beta_I}{N}\mathbf{I}_{NN}(t_j)\mathbf{S}_{NN}(t_j)\right|^2 \\
& +\frac{1}{N_f}\sum_{j=1}^{N_f}\left|\frac{(\Delta t)^{\kappa_{3_{NN}}(t_j)-1}}{\Gamma(\kappa_{3_{NN}}(t_j)+1)}\delta_t^{\kappa_{3_{NN}}(t_j)}\mathbf{H}^c_{NN}(t_j)-p_{NN}(t_j)\gamma\mathbf{I}_{NN}(t_j)\right|^2 \\
& +\frac{1}{N_f}\sum_{j=1}^{N_f}\left|N-\mathbf{S}_{NN}(t)-\mathbf{I}_{NN}(t)-\mathbf{H}_{NN}(t)-\mathbf{D}_{NN}(t)-\mathbf{R}_{NN}(t)\right|^2.
\end{aligned}
\tag{17}
$$

Detailed expansions of the ODE residual for model $\mathbb{F}_2$ can be similarly derived.

#### 5.5 Reducing Computational Complexity of fPINNs

The computational bottleneck in the fractional models is the for loop in computing the fractional derivative. It becomes more challenging when we decrease the time step $\tau$. Here, we show a vectorization technique that we implemented to reduce the computational cost in obtaining the fractional differentiation matrix related to variable-order Caputo derivative. Thus, we have

$$\begin{bmatrix} \delta_t^{\kappa_0} f_0 \\ \delta_t^{\kappa_1} f_1 \\ \delta_t^{\kappa_2} f_2 \\ \vdots \\ \delta_t^{\kappa_{M-1}} f_{M-1} \\ \delta_t^{\kappa_M} f_M \end{bmatrix} = \begin{bmatrix} \frac{\tau^{-\kappa_0}}{\Gamma(2-\kappa_0)} & \frac{\tau^{-\kappa_0}}{\Gamma(2-\kappa_0)} & \cdots & \frac{\tau^{-\kappa_0}}{\Gamma(2-\kappa_0)} \\ \frac{\tau^{-\kappa_1}}{\Gamma(2-\kappa_1)} & \frac{\tau^{-\kappa_1}}{\Gamma(2-\kappa_1)} & \cdots & \frac{\tau^{-\kappa_1}}{\Gamma(2-\kappa_1)} \\ \frac{\tau^{-\kappa_2}}{\Gamma(2-\kappa_2)} & \frac{\tau^{-\kappa_2}}{\Gamma(2-\kappa_2)} & \cdots & \frac{\tau^{-\kappa_2}}{\Gamma(2-\kappa_2)} \\ \vdots & \vdots & \cdots & \vdots \\ \frac{\tau^{-\kappa_{M-1}}}{\Gamma(2-\kappa_{M-1})} & \frac{\tau^{-\kappa_{M-1}}}{\Gamma(2-\kappa_{M-1})} & \cdots & \frac{\tau^{-\kappa_{M-1}}}{\Gamma(2-\kappa_{M-1})} \\ \frac{\tau^{-\kappa_M}}{\Gamma(2-\kappa_M)} & \frac{\tau^{-\kappa_M}}{\Gamma(2-\kappa_M)} & \cdots & \frac{\tau^{-\kappa_M}}{\Gamma(2-\kappa_M)} \end{bmatrix} \odot \mathbf{W} \times \begin{bmatrix} f_0 \\ f_1 \\ f_2 \\ \vdots \\ f_{M-1} \\ f_M \end{bmatrix} \tag{18}$$

where $\odot$ and $\times$ denote the Hadamard product and matrix multiplication, respectively. The coefficient matrix $\mathbf{W}$ can be obtained by $\mathbf{W} = [\mathbf{W}_0 ** \mathbf{C} - \mathbf{W}_1 ** \mathbf{C}] + [\mathbf{W}_2 ** \mathbf{C} - 2\mathbf{W}_3 ** \mathbf{C} + \mathbf{W}_4 ** \mathbf{C}] + \bar{\mathbf{I}}$, where $**$ denotes point-wise power operator and the matrices on the right-hand side are given by

$$\mathbf{C} = \begin{bmatrix} 1-\kappa_0 & 1-\kappa_0 & 1-\kappa_0 & \cdots & 1-\kappa_0 & 1-\kappa_0 \\ 1-\kappa_1 & 1-\kappa_1 & 1-\kappa_1 & \cdots & 1-\kappa_1 & 1-\kappa_1 \\ 1-\kappa_2 & 1-\kappa_2 & 1-\kappa_2 & \cdots & 1-\kappa_2 & 1-\kappa_2 \\ \vdots & \vdots & \vdots & \cdots & \vdots & \vdots \\ 1-\kappa_{M-1} & 1-\kappa_{M-1} & 1-\kappa_{M-1} & \cdots & 1-\kappa_{M-1} & 1-\kappa_{M-1} \\ 1-\kappa_M & 1-\kappa_M & 1-\kappa_M & \cdots & 1-\kappa_M & 1-\kappa_M \end{bmatrix}, \tag{19}$$

$$\bar{\mathbf{I}} = \begin{bmatrix} 0 & 0 & 0 & \cdots & 0 & 0 \\ 0 & 1 & 0 & \cdots & 0 & 0 \\ 0 & 0 & 1 & \cdots & 0 & 0 \\ \vdots & \vdots & \vdots & \ddots & \vdots & \vdots \\ 0 & 0 & 0 & \cdots & 1 & 0 \\ 0 & 0 & 0 & \cdots & 0 & 1 \end{bmatrix}, \quad \mathbf{W}_0 = \begin{bmatrix} 0 & 0 & 0 & \cdots & 0 & 0 \\ 0 & 0 & 0 & \cdots & 0 & 0 \\ 1 & 0 & 0 & \cdots & 0 & 0 \\ \vdots & \vdots & \vdots & \ddots & \vdots & \vdots \\ (M-2) & 0 & 0 & \cdots & 0 & 0 \\ (M-1) & 0 & 0 & \cdots & 0 & 0 \end{bmatrix}, \tag{20}$$

$$\mathbf{W}_1 = \begin{bmatrix} 0 & 0 & 0 & \cdots & 0 & 0 \\ 1 & 0 & 0 & \cdots & 0 & 0 \\ 2 & 0 & 0 & \cdots & 0 & 0 \\ \vdots & \vdots & \vdots & \ddots & \vdots & \vdots \\ (M-1) & 0 & 0 & \cdots & 0 & 0 \\ M & 0 & 0 & \cdots & 0 & 0 \end{bmatrix}, \quad \mathbf{W}_2 = \begin{bmatrix} 0 & 0 & 0 & \cdots & 0 & 0 \\ 0 & 0 & 0 & \cdots & 0 & 0 \\ 0 & 2 & 0 & \cdots & 0 & 0 \\ \vdots & \vdots & \vdots & \cdots & \vdots & \vdots \\ 0 & (M-1) & (M-2) & \cdots & 1 & 0 \\ 0 & M & (M-1) & \cdots & 2 & 0 \end{bmatrix}, \tag{21}$$

$$\mathbf{W}_3 \begin{bmatrix} 0 & 0 & 0 & \cdots & 0 & 0 \\ 0 & 0 & 0 & \cdots & 0 & 0 \\ 0 & 1 & 0 & \cdots & 0 & 0 \\ \vdots & \vdots & \vdots & \cdots & \vdots & \vdots \\ 0 & (M-2) & (M-3) & \cdots & 1 & 0 \\ 0 & (M-1) & (M-2) & \cdots & 1 & 0 \end{bmatrix}, \quad \mathbf{W}_4 \begin{bmatrix} 0 & 0 & 0 & \cdots & 0 & 0 \\ 0 & 0 & 0 & \cdots & 0 & 0 \\ 0 & 0 & 0 & \cdots & 0 & 0 \\ \vdots & \vdots & \vdots & \cdots & \vdots & \vdots \\ 0 & (M-3) & (M-4) & \cdots & 1 & 0 \\ 0 & (M-2) & (M-3) & \cdots & 0 & 0 \end{bmatrix}. \tag{22}$$
